# Polygenic Hazard Score for Predicting Age-associated Risk of Alzheimer’s Disease in European Populations: Development and Validation

**DOI:** 10.1101/2025.07.28.25332293

**Authors:** Bayram Cevdet Akdeniz, Shahram Bahrami, Espen Hagen, Julian Fuhrer, Vera Fominykh, Alexey Shadrin, Tahir Tekin Filiz, Lavinia Athanasiu, Benjamin Grenier-Boley, Céline Bellenguez, Itziar de Rojas, Fahri Küçükali, Anja Schneider, Luca Kleineidam, Dan Rujescu, Norbert Scherbaum, Jürgen Deckert, Steffi Riedel-Heller, Lucrezia Hausner, Laura Molina-Porcel, Timo Grimmer, Stefanie Heilmann-Heimbach, Susanne Moebus, Nikolaos Scarmeas, Jose María García-Alberca, Emilio Franco-Macías, Pablo Mir, Luis M. Real, Eloy Rodriguez-Rodriguez, Jose Luís Royo, María Eugenia Sáez, Ángel Carracedo, Adolfo Lopez de Munain, Guillermo Amer-Ferrer, Miguel Calero, Miguel Medina, Guillermo Garcia-Ribas, Maite Mendioroz, Oriol Dols-Icardo, Fermin Moreno, Jordi Pérez-Tur, María J. Bullido, Victoria Álvarez, Hilkka Soininen, Sami Heikkinen, Alexandre de Mendonça, Shima Mehrabian, Latchezar Traykov, Jakub Hort, Martin Vyhnalek, Nicolai Sandau, Jesper Qvist Thomassen, Jiao Luo, Yolande A.L. Pijnenburg, Niccolo Tesi, John van Swieten, Vilmantas Giedraitis, Julie Williams, Gael Nicolas, Stephanie Debette, Philippe Amouyel, Edna Grünblatt, Julius Popp, Paola Bossù, Daniela Galimberti, Giacomina Rossi, Beatrice Arosio, Patrizia Mecocci, Alessio Squassina, Lucio Tremolizzo, Barbara Borroni, Benedetta Nacmias, Davide Seripa, Innocenzo Rainero, Antonio Daniele, Fabrizio Piras, Carlo Masullo, Patrick Gavin Kehoe, Ruth Frikke-Schmidt, Roberta Ghidoni, Agustín Ruiz, Victoria Fernandez, Pascual Sánchez-Juan, Kristel Sleegers, Martin Ingelsson, Mikko Hiltunen, Rebecca Sims, Alfredo Ramirez, Iris J. Broce, Jan Haavik, Geir Selbæk, Anne-Brita Knapskog, Ingvild Saltvedt, Sverre Bergh, Eivind Aakhus, Bjørn-Eivind Kirsebom, Leiv Otto Watne, Arvid Rongve, Dag Årsland, Srdjan Djurovic, Eystein Stordal, Mathias Toft, Katja Scheffler, Tormod Fladby, Jean-Charles Lambert, Anders Dale, Oleksandr Frei, Ole Andreassen

**Affiliations:** Centre for Precision Psychiatry, Division of Mental Health and Addiction, Oslo University Hospital & Institute of Clinical Medicine, University of Oslo, Oslo, Norway; Department of Pharmacy, Section for Pharmacology and Pharmaceutical Biosicences, University of Oslo, Oslo, Norway; Univ. Lille, Inserm, CHU Lille, Institut Pasteur de Lille, LabEx DISTALZ - U1167-RID-AGE Facteurs de risque et déterminants moléculaires des maladies liées au vieillissement, Lille, France; Ace Alzheimer Center Barcelona, Universitat Internacional de Catalunya (UIC), Barcelona, Spain; Networking Research Center on Neurodegenerative Diseases (CIBERNED), Instituto de Salud Carlos III, Madrid, Spain; Luxembourg Centre for Systems Biomedicine (LCSB), University of Luxembourg, Luxembourg; Complex Genetics of Alzheimer’s Disease Group, VIB Center for Molecular Neurology, VIB, Antwerp, Belgium; Department of Biomedical Sciences, University of Antwerp, Antwerp, Belgium; German Center for Neurodegenerative Diseases (DZNE), Bonn, Germany; Department of Old Age Psychiatry and Cognitive Disorders, University Hospital Bonn, Venusberg-Campus 1, 53127 Bonn, Germany; Department of Psychiatry and Psychotherapy, Medical University of Vienna, Vienna, Austria; Comprehensive Center for Clinical Neurosciences and Mental Health, Medical University of Vienna, Vienna, Austria; Department of Psychiatry and Psychotherapy, LVR-University Hospital Essen, Medical Faculty of the University of Duisburg-Essen, Essen, Germany; Department of Psychiatry, Psychosomatics and Psychotherapy, Center of Mental Health, University Hospital of Würzburg, Germany; Institute of Clinical Epidemiology and Biometrics, Julius-Maximilians-University Würzburg, Würzburg, Germany; Institute of Social Medicine, Occupational Health and Public Health, University of Leipzig, 04103 Leipzig, Germany; Department of Geriatric Psychiatry, Central Institute for Mental Health Mannheim, Faculty Mannheim, University of Heidelberg, Germany; Neurological Tissue Bank - Biobanc-Hospital Clinic -IDIBAPS, Barcelona, Spain; Alzheimer’s disease and other cognitive disorders Unit, Neurology Department, Hospital Clinic, Barcelona, Spain; CIBERNED, Network Center for Biomedical Research in Neurodegenerative Diseases, National Institute of Health Carlos III, Madrid, Spain; Center for Cognitive Disorders, Department of Psychiatry and Psychotherapy, TUM University Hospital, Technical University of Munich, School of Medicine, Munich, Germany; Institute of Human Genetics, University of Bonn, School of Medicine & University Hospital Bonn, Bonn, Germany; Institute for Urban Public Health, University Hospital of University Duisburg-Essen, Essen, Germany; Taub Institute for Research in Alzheimer’s Disease and the Aging Brain, The Gertrude H. Sergievsky Center, Department of Neurology, Columbia University, New York, NY, USA; 1st Department of Neurology, Aiginition Hospital, National and Kapodistrian University of Athens, Medical School, Athens, Greece; Alzheimer Research Center & Memory Clinic, Instituto Andaluz de Neurociencia (IANEC), Málaga, Spain; Unidad de Demencias, Servicio de Neurología, Instituto de Biomedicina de Sevilla (IBiS), Hospital Universitario Virgen del Rocío/CSIC/Universidad de Sevilla, Seville, Spain; Unidad de Trastornos del Movimiento, Servicio de Neurología, Instituto de Biomedicina de Sevilla, Hospital Universitario Virgen del Rocío/CSIC/Universidad de Sevilla, Seville, Spain; Departamento de Medicina, Facultad de Medicina, Universidad de Sevilla, Seville, Spain; Unidad Clínica de Enfermedades Infecciosas y Microbiología, Hospital Universitario de Valme, Sevilla, Spain; Departamento de Bioquímica Médica, Biología Molecular e Inmunología, Facultad de Medicina, Universidad de Sevilla, Sevilla, Spain; CIBERINFEC, Network Center for Biomedical Research in Infectious Diseases, National Institute of Health Carlos III, Madrid, Spain; Neurology Service, Marqués de Valdecilla University Hospital (University of Cantabria and IDIVAL), Santander, Spain; Departamento de Medicina y Psiquiatría, Universidad de Cantabria, Santander, Spain; Departamento de Especialidades Quirúrgicas, Bioquímica e Inmunología, Facultad de Medicina, Universidad de Málaga, Málaga, Spain; CAEBI, Centro Andaluz de Estudios Bioinformáticos, Sevilla, Spain; Grupo de Medicina Xenómica, Centro en Red de Investigación en Enfermedades Raras (CIBERER-ISCIII), CIMUS, Universidade de Santiago de Compostela, Santiago de Compostela, Spain; Fundación de Medicina Xenómica, Servicio Galego de Saúde (SERGAS), Instituto de Investigación Sanitaria de Santiago (IDIS), Santiago de Compostela, Spain; Department of Neurology, Hospital Universitario Donostia-Instituto Biodonostia-OSAKIDETZA-EHU-CIBERNED, San Sebastian, Spain; Department of Neurosciences, Faculty of Medicine and Nursery, University of the Basque Country, San Sebastián, Spain; Department of Neurology, Hospital Universitario Son Espases, Palma, Spain; Chronic Disease Programme (UFIEC), Instituto de Salud Carlos III, Madrid, Spain; CIEN Foundation/Queen Sofia Foundation Alzheimer Center, Madrid, Spain; Hospital Universitario Ramon y Cajal, IRYCIS, Madrid, Spain; Navarrabiomed, Pamplona, Spain; Department of Neurology, Hospital Universitario de Navarra, Pamplona, Spain; Sant Pau Memory Unit, IR SANT PAU, Hospital de la Santa Creu i Sant Pau, Barcelona, Spain; Biogipuzkoa Health Research Institute, Neurosciences Area, Group of Neurodegenerative Diseases, 20014 San Sebastián, Spain; Unitat de Genètica Molecular, Institut de Biomedicina de València-CSIC, Valencia, Spain; Centro de Biología Molecular Severo Ochoa (UAM-CSIC), Madrid, Spain; Instituto de Investigacion Sanitaria ‘Hospital la Paz’ (IdIPaz), Madrid, Spain; Universidad Autónoma de Madrid, Madrid, Spain; Laboratorio de Genética, Hospital Universitario Central de Asturias, Oviedo, Spain; Instituto de Investigación Sanitaria del Principado de Asturias (ISPA), Oviedo, Spain; Institute of Clinical Medicine - Neurology, University of Eastern Finland, Kuopio, Finland; Institute of Biomedicine, University of Eastern Finland, Kuopio, Finland; Faculty of Medicine, University of Lisbon, Lisbon, Portugal; Clinic of Neurology, UH “Alexandrovska”, Medical University - Sofia, Sofia, Bulgaria; Memory Clinic, Department of Neurology, Charles University, Second Faculty of Medicine and Motol University Hospital, Prague, Czech Republic; International Clinical Research Center, St. Anne’s University Hospital Brno, Brno, Czech Republic; Department of Clinical Biochemistry, Copenhagen University Hospital - Rigshospitalet, Copenhagen, Denmark; Alzheimer Center Amsterdam, Department of Neurology, Amsterdam Neuroscience, Vrije Universiteit Amsterdam, Amsterdam UMC, Amsterdam, The Netherlands; Delft Bioinformatics Lab, Delft University of technology, Delft, The Netherlands; Department of Neurology, ErasmusMC, Rotterdam, The Netherlands; Dept. of Public Health and Caring Sciences / Geriatrics, Uppsala University, Uppsala, Sweden; UKDRI@ Cardiff, School of Medicine, Cardiff University, Cardiff, UK; Centre for Neuropsychiatric Genetics and Genomics, Division of Psychological Medicine and Clinical Neuroscience, School of Medicine, Cardiff University, Cardiff, UK; Univ Rouen Normandie, Normandie Univ, Inserm U1245 and CHU Rouen, Department of Genetics and CNR-MAJ, F-76000 Rouen, France; University of Bordeaux, Inserm Bordeaux Population Health Research Center U1219, Bordeaux, France; CHU de Bordeaux, Pole santé publique, Bordeaux, France; Institut du Cerveau (ICM), Paris Brain Institute, INSERM U1127, UMR CNRS 7225 Paris, Sorbonne Université, Assistance Publique des Hôpitaux de Paris, Paris, France; Department of Child and Adolescent Psychiatry and Psychotherapy, University Hospital of Psychiatry Zurich, University of Zurich, Zurich, Switzerland; Neuroscience Center Zurich, University of Zurich and ETH Zurich, Zurich, Switzerland; Zurich Center for Integrative Human Physiology, University of Zurich, Zurich, Switzerland; Old Age Psychiatry, Department of Psychiatry, Lausanne University Hospital, Lausanne, Switzerland; Department of Adult Psychiatry, University Hospital of Psychiatry Zürich, Zürich, Switzerland; Institute for Regenerative Medicine, University of Zürich, Zürich, Switzerland; Experimental Neuro-psychobiology Laboratory, IRCCS Fondazione Santa Lucia, Rome, Italy; Neurodegenerative Diseases Unit, Fondazione IRCCS Ca’ Granda, Ospedale Policlinico, Milan, Italy; Dept. of Biomedical, Surgical and Dental Sciences, University of Milan, Milan, Italy; Unit of Neurology 8, Fondazione IRCCS Istituto Neurologico Carlo Besta, Milan, Italy; Department of Clinical Sciences and Community Health, University of Milan, 20122 Milan, Italy; Geriatric Unit, Fondazione IRCCS Ca’ Granda Ospedale Maggiore Policlinico, 20122 Milan, Italy; Institute of Gerontology and Geriatrics, Department of Medicine and Surgery, University of Perugia, Perugia, Italy; Division of Clinical Geriatrics, Department of Neurobiology, Care Sciences and Society, Karolinska Institutet, Stockholm, Sweden; Department of Biomedical Sciences, Section of Neuroscience and Clinical Pharmacology, University of Cagliari, Cagliari, Italy; Neurology Unit, IRCCS “San Gerardo dei Tintori”, Monza, Italy; School of Medicine and Surgery, University of Milano-Bicocca, Monza, Italy; Department of Clinical and Experimental Sciences, University of Brescia, Brescia, Italy; Molecular Markers Laboratory, IRCCS Istituto Centro San Giovanni di Dio Fatebenefratelli, Brescia, Italy; Department of Neuroscience, Psychology, Drug Research and Child Health, University of Florence, Florence, Italy; IRCCS Fondazione Don Carlo Gnocchi, Florence, Italy; Department of Hematology and Stem Cell Transplant, Vito Fazzi Hospital, Lecce, Italy; Department of Neuroscience “Rita Levi Montalcini”, University of Torino, Torino, Italy; Department of Neuroscience, Università Cattolica del Sacro Cuore, Rome, Italy; Neurology Unit, IRCCS Fondazione Policlinico Universitario A. Gemelli, Rome, Italy; Laboratory of Neuropsychiatry, IRCCS Santa Lucia Foundation, Rome, Italy; Institute of Neurology, Catholic University of the Sacred Heart, Rome, Italy; Translational Health Sciences, Bristol Medical School, University of Bristol, Bristol, UK; Department of Clinical Medicine, University of Copenhagen, Copenhagen, Denmark; Biggs Institute for Alzheimer’s and Neurodegenerative Diseases, University of Texas Health Science Center, San Antonio, TX, USA; Department of Microbiology, Immunology, and Molecular Genetics, University of Texas Health San Antonio, TX 78229-3900, USA; Alzheimer’s Centre Reina Sofia-CIEN Foundation-ISCIII, Madrid, Spain; Krembil Brain Institute, University Health Network, Toronto, Ontario, Canada; Tanz Centre for Research in Neurodegenerative Diseases, Departments of Medicine and Laboratory Medicine & Pathobiology, University of Toronto, Toronto, Ontario, Canada; Division of Neurogenetics and Molecular Psychiatry, Department of Psychiatry and Psychotherapy, Faculty of Medicine and University Hospital Cologne, University of Cologne, Cologne, Germany; Department of Psychiatry & Glenn Biggs Institute for Alzheimer’s and Neurodegenerative Diseases, San Antonio, TX, USA; Cologne Excellence Cluster on Cellular Stress Responses in Aging-Associated Disease (CECAD), University of Cologne, Cologne, Germany; Center for Multimodal Imaging and Genetics, University of California San Diego, La Jolla, CA, USA; Memory and Aging Center, Department of Neurology, Weill Institute for Neurosciences, University of California San Francisco, San Francisco, CA, USA; Department of Biomedicine, University of Bergen, Bergen, Norway; Bergen Center for Brain Plasticity, Division of Psychiatry, Haukeland University Hospital, Bergen, Norway; Faculty of Medicine, University of Oslo, Oslo, Norway; Department of Geriatric Medicine, Oslo University Hospital, Oslo, Norway; Norwegian National Centre for Ageing and Health, Vestfold Hospital Trust, Tønsberg, Norway; Department of Geriatrics, St. Olav’s Hospital, Trondheim University Hospital, Trondheim, Norway and Department of Neuromedicine and Movement Science, Norwegian University of Science and Technology (NTNU), Trondheim, Norway; Research Centre for Age-related Functional Decline and Disease, Innlandet Hospital Trust, Ottestad, Norway; Department of Neurology, University Hospital of North Norway, Tromsø; Department of Psychology, Faculty of Health Sciences, UiT The Arctic University of Norway, Tromsø, Norway; Oslo Delirium Research Group, Department of Geriatric Medicine, Akershus University Hospital, Lørenskog, Norway; Department of Research and Innovation, Helse Fonna, Haugesund, Norway; University of Bergen, Department of Clinical Medicine (K1), Bergen, Norway; Centre for Age-Related Medicine, Stavanger University Hospital, Stavanger, Norway; Department of Old Age Psychiatry, Institute of Psychiatry, Psychology and Neuroscience, King’s College London, London, UK; Department of Medical Genetics, Division of Mental Health and Addiction, Oslo University Hospital, Oslo, Norway; Department of Psychiatry, Namsos Hospital, Namsos, Norway; Department of Neurology, Oslo University Hospital, Oslo, Norway; Department of Neurology and Clinical Neurophysiology, University Hospital of Trondheim, Trondheim, Norway; Department of Neuromedicine and Movement Science, Norwegian University of Science and Technology, Trondheim, Norway; Department of Neurology, Akershus University Hospital, Lørenskog, Norway; Department of Radiology, School of Medicine, University of California San Diego, La Jolla, CA, USA; Department of Cognitive Science, University of California San Diego, La Jolla, CA, USA; Institute of Clinical Medicine, University of Oslo, Oslo, Norway

## Abstract

**Objectives:** Polygenic hazard score (PHS) models can be used to predict the age-associated risk for complex diseases, including Alzheimer’s disease (AD). In this study, we present an improved PHS model for AD that incorporates a large number of genetic variants and demonstrates enhanced predictive accuracy for age of onset in European populations compared to alternative models.

**Methods:** We used the genotyped European Alzheimer & Dementia Biobank (EADB) sample (n=42,120) to develop and evaluate the performance of the PHS model. We developed a PHS model building on 720 genetic variants, including Apolipoprotein E (*APOE*) ε2 and ε4 alleles. We used Elastic Net-regularized Cox regression approach to develop the PHS model.

**Results:** The new PHS model (EADB720) improved prediction accuracy compared to alternative models in European populations, with the Odds Ratio OR80/20 from the highest quintile of risk (80th risk percentile and above) to the lowest quintile of risk (20th risk percentile and below) varying between 5.10 and 13.15 within the range of age of onset from 65 – 85 years. Our model also improved risk stratification across ε3/3 individuals of European ancestry (OR80/20 ranges from 1.95 to 3.52). It was also successfully validated in independent datasets (HUSK, DemGene and ADNI) by achieving OR80/20 up to 10.00 in each independent dataset.

**Conclusion:** Our EADB720 model significantly improves the accuracy of age-associated risk of AD across European populations (pval<0.03). Accurately predicting the age of onset of AD is of large clinical importance to implementing new AD medication and early intervention in clinical settings.

## Introduction

Late-onset Alzheimer’s disease (AD) is the most common form of dementia and typically manifests after the age of 65. It is estimated that around 13% of people above the age of 65 and 45% over the age of 85 have AD [1],[2] making it a global health challenge. Heritability for AD is high, with genetic factors accounting for approximately 60-80% of the total risk [3]. These genetic variants include the apolipoprotein E (*APOE*) ε4 and ε2 alleles [1], [4] and various other single nucleotide polymorphisms (SNPs) [5],[6].

The prevalence of AD continues to rise as the population ages, posing substantial challenges to public health systems worldwide. Prediction of AD onset is crucial for several reasons. The pathobiological changes underlying AD may begin up to 20 years before the onset of clinical symptoms [7],[8]. Identifying individuals at risk before significant cognitive decline occurs allows for timely intervention strategies that can slow or possibly prevent the onset of the disease, potentially preserving quality of life and reducing the burden on caregivers and healthcare systems [9], [10].

Early prediction also enables the stratification of patients in clinical trials for emerging therapies, contributing to advancing treatment options. Importantly, it allows for the selection of patients for the new monoclonal antibody medications currently in the pipeline [11],[12]. The risk of Amyloid-related imaging abnormalities (ARIA) changes based on the choice of the anti-amyloid medications and the number of ε4 copies that a patient carries [13]. Thus, developing a reliable risk stratification model for carriers and non-carriers of ε4 is critical for informed drug selection. Moreover, understanding individual risk can facilitate proactive lifestyle changes and planning for future care needs, ultimately enhancing patient outcomes and reducing the societal burden of this devastating condition [7].

Since AD is highly heritable [14], using genetic data is relevant to predict the risk of developing AD. Polygenic Hazard Score (PHS) is a Cox proportional hazards model for predicting the age-related risk of developing a disease. The Desikan PHS model for AD [15] included 33 SNPs and *APOE* variants (ε2 and ε4), developed using International Genomics of Alzheimer’s Project (IGAP) samples and validated Alzheimer’s Disease Genetics Consortium [15], and subsequently validated in different populations [16], [17]. However, the Desikan PHS model has not been validated for the cohorts from different European regions. Furthermore, the Desikan model was developed based on a stepwise procedure for variant selection, while recent studies show that regularized approaches (LASSO, Ridge, ElasticNet) perform better for Cox’s proportional hazards models [18], [19].

Here, we developed an improved AD PHS model using the European Alzheimer & Dementia Biobank (EADB) dataset, additionally informing our selection of genetic variants by the largest GWAS studies available today. ElasticNet-regularized Cox regression resulted in a PHS model with 720 SNPs (EADB720) that performs significantly better than the Desikan PHS model in European samples. We incorporated model training using cross-validation and leave-one-country-out analysis within EADB and tested the final model in three independent cohorts (HUSK, DEMGENE, ADNI).

## Methods

### Participant Samples

#### European Alzheimer & Dementia Biobank (EADB)

EADB dataset consists of 20,301 AD cases and 21,839 controls from 15 European countries (Belgium, Bulgaria, Czech Republic, Denmark, Finland, France, Germany, Greece, Italy, Portugal, Spain, Sweden, Switzerland, the Netherlands and Great Britain). Genotypes are available for all N=42,140 subjects at approx. 117 million variants after TOPMed imputation (after filtering on R2≥0.3 imputation score from minimac4). We applied a basic QC procedure for SNP selection with the following parameters using PLINK [20] (--maf 0.005 --hwe 1e-60 --geno 0.02), leading to 7,622,793 SNPs considered for further analysis.

Phenotype data included AD status as a binary variable, participant’s age at baseline, age of onset for cases, and age of last examination for controls. To derive the time-to-event variable (in years) for Cox regression model, we used age of onset for cases and age at last examination for controls. If age of last examination was missing for controls, then age at baseline was used instead. As our focus was on late-onset AD, we filtered out cases with age at onset below 60 and controls with age at last follow-up below 60 as applied in [15]. Additionally, we excluded 466 IGAP-related subjects who were included in the development of the Desikan PHS model (part of the GERAD study). Furthermore, we excluded subjects due to missing information on age of onset, age of last follow-up, or *APOE* status. This resulted in a final sample of 13,604 cases, 16,608 controls as summarized in Table 1 (see Supplementary Fig. 1 for further details). More details regarding the dataset can be obtained from [21].

**Table 1.** EADB cohort and *APOE* status.

|  | Cases | Controls |
| --- | --- | --- |
| Total Sample (n) | 13,604 | 16,608 |
| Female (%) | 62.8 | 56.2 |
| Mean (and standard deviation) of age of onset for and age of last follow up for controls | 74.3<br>(7.71) | 72.5<br>(7.62) |
| $\epsilon 2/2$ (%) | 1.3 | 0.7 |
| $\epsilon 2/3$ (%) | 4.3 | 11.4 |
| $\epsilon 3/3$ (%) | 40.1 | 63.5 |
| $\epsilon 3/4$ (%) | 41.1 | 20.9 |
| $\epsilon 2/4$ (%) | 2.2 | 1.8 |
| $\epsilon 4/4$ (%) | 11.0 | 1.7 |

#### The Hordaland Health Studies (HUSK)

HUSK is a population-based cohort from Hordaland, Norway. Approximately 36,000 residents of Hordaland County participated, with about 18,000 in the 1992/93 survey and around 26,000 in the 1997/99 survey. Nearly all participants invited in 1992-93 were born in 1925-27 and 1950-52, with a few people born in 1928-57. The participants invited in 1997-99 were born in 1953-57. About 7,000 individuals participated in both surveys (https://husk-en.w.uib.no/).

HUSK participants were genotyped at deCODE Genetics (https://www.decode.com/) in two batches (10,083 and 25,432 individuals in batch 1 and 2 respectively) using a customized Illumina GSA v3 array. After performing QC and imputation to genotype data (See Supplementary Notes for details) the resulting dataset contained 10,531,075 variants and 35,476 individuals.

Phenotype data for the HUSK sample were obtained from the Norwegian Patient Registry (NPR) and death-year registry. We used ICD10 codes (G30, F00 and all subcodes) to determine AD cases. For controls we excluded samples with any AD; cognitive impairment (F70-F79, F80-89, F99 with all subcodes); dementia (F01, F02, F03, F04, F05.1, F06 and all subcodes); and all neurodegenerative disorders with CNS involvement (G10-G13, G20-G23, G31-G32 and all subcodes). For cases, age of onset is determined by using “year_of_first_diag_AD” while for controls “year_last_visit” and death-year registry information were used for age of last follow-up (for details see Supplementary Table S2a).

Applying the same phenotyping filtering procedure with respect to age as done in the EADB dataset (age≥60) on HUSK, we used 757 AD cases for validation. (for details see Supplementary Table S2a). For controls, as described above, we kept patients with non-AD related diseases, resulting in 24,719 samples. We conducted the validation both with and without controls under the permission of REC-vest (permission #2018/915).

#### DemGene

The DemGene cohort was recruited from a network of clinical sites across Norway that systematically gather data through standardized assessments of cognitive, functional, and behavioral measures from real-world patients in Norway included in The Norwegian Registry of Persons Assessed for Cognitive Symptoms (NorCog) [22] and and Dementia Disease Initiation (DDI) [23]. Participants in DemGene were genotyped at various genotyping centers with the majority processed at deCODE Genetics. Genotyping was performed in 28 batches using different genotyping arrays, most commonly Illumina GSA v1 or v3, as well as customized versions of these arrays, Illumina Human OmniExpress-12 and OmniExpress-24, Affymetrix 6.0. For this study, we included only Norwegian-origin cohorts for validation purposes.

After performing quality control (QC) and imputation of DemGene data (See Supplementary Notes for details), and filtering based on the Norwegian-origin samples, AD cases were identified based on the presence of diagnostic codes for AD, AD-Mild Cognitive Impairment (AD-MCI), AD-Subjective cognitive decline (AD-SCD) or confirmed amyloid-positive. There are three different variables related to age of onset:

1. “demo_age_diagnosis”, ii) “demo_age_onset”, iii) “demo_age_symptom_start”. When some of these variables were missing, we used them in a predefined priority order. For controls, we used “demo_age” variable for age of last follow-up (for details see Supplementary Table S2b). After applying the same exclusion procedure as done for EADB, we used 2137 AD cases and 1371 controls for the independent testing of our approaches with REC# 2014/631.

#### Alzheimer’s Disease Neuroimaging Initiative (ADNI) Study

For validation, we also used data from Alzheimer’s Disease Neuroimaging Initiative (ADNI) study, including N=830 participants with dementia diagnosis (cases), and N=1600 participants without any dementia diagnosis (controls). For cases, age of onset was defined as their age at the earliest visit where they received diagnosis. For controls, censoring age was defined based on last visit. Imputation was performed across ADNI1/2/3/GO subsets of ADNI using TOPMed Imputation Server, with final imputed datasets including 11,377,441 variants for 2027 subjects, covering 712 cases and 1315 controls.

#### Genome-wide association studies (GWAS) summary statistics

List of external GWAS studies that we used for meta-analysis is listed as follows: IGAP (17,008 cases and 37,154 controls) [24], FinnGen consortium (9,271 cases and 299,883 controls) [25], and Psychiatric Genomic Consortium (PGC) UK Biobank (UKB) AD [6] (46,613 proxy cases and 318,246 controls). We performed meta-analysis on these summary statistics as will be defined in the following section.

### Statistical Approach

Our statistical approach for developing the PHS model for AD involves 3 steps: (1) selecting candidate SNPs for inclusion in the PHS model from an external GWAS; (2) performing variable selection using Elastic Net-based Cox regression for PHS model development and (3) evaluate the performance of the developed PHS model.

#### 1) Selection of Candidate SNPs

Selecting candidate SNPs for inclusion into our PHS model was based on performing a meta-analysis on GWAS data from IGAP, FinnGen and PGC UKB. The meta-analysis was performed in two stages. As the UKB GWAS is based on proxy AD cases: first we performed the meta-analysis for all cohorts except UKB. Second, we meta-analyzed the output from step 1 with the UKB data. This meta-analysis was sample size weighted, whereas step 1 was variance-based. Further, we used Neffective as a weight in step 1 and N/4 as a weight for UKB in step 2. Neffective is the sum across all datasets for that variant (4/((1/Ncases)+(1/Ncontrols)), adjusting for case-control imbalance. The UKB Neffective is defined as N/4 to adjust for the dilute nature of the proxy phenotype.

Using the cleansumstats v1.7.0 pipeline [26], indels, ambiguous A/T and C/G SNPs, SNPs without rs# in dbSNP b156 were removed, leading to 7,622,793 SNPs after QC procedure, of which 7,571,993 SNPs were shared between GWAS meta-analysis and EADB genotype data. 7247 SNPs with pval<1e-5 in the GWAS meta-analysis were included for variable selection in the PHS model training step.

#### 2) ElasticNet-regularized PHS model

The EADB individual-level dataset was used for model development 80% for training, including nested cross-validation for optimizing parameters of ElasticNet regularization and 20% for validation. This procedure was repeated 100 times to obtain the mean and standard deviation estimates of the performance metrics described in step 3. Using the training dataset, we developed a PHS model by selecting variables among 7247 candidate variants prefiltered from the meta-analysis. The model development, i.e., determining the effect size of the variants (𝑏_!_) was done using a machine-learning ElasticNet-regularized approach via Cox regression [27].

Cox regression requires a time-to-event variable and a censoring variable. Censoring allows for the inclusion of subjects who have not experienced the event of interest by the end of the study or when a subject is no longer followed up on before experiencing the event (i.e., AD onset). The censoring variable is coded as a binary variable, where 1 is used to denote an event (AD onset, cases), and 0 is used to denote censoring (AD free, controls). Cox regression was applied through the GLMNET package [28]. The effect sizes of the variants were determined by maximizing the partial likelihood function of the Cox model (ℓ(β)) with penalty terms defined as:

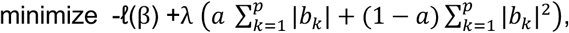

where λ is the tuning parameter controlling the overall strength of the penalty term and 𝑎 is controlling term between LASSO (𝑎 = 1) and Ridge (𝑎 = 0) regression (and for ElasticNet 0<a<1). The optimal tuning parameters (λ,𝑎) were determined by performing 5-fold cross-validation on training data. We also controlled country status during model development by including this variable (without penalizing it) during the development and excluding the effect size of country status in validation.

Having determined the optimal tuning parameter of the ElasticNet, we then developed the model by determining the effect sizes of the SNPs (𝑏_!_) on training data. Finally, we applied this developed model on the 20% validation data within EADB. The PHS of individual *i* (𝑃𝐻𝑆_&_) was calculated using the corresponding genotype vector (𝑋_&_) and the PHS model’s effect sizes (𝑏_!_) as

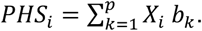

For validation of the external datasets, we used the whole EADB dataset to develop the ultimate model (EADB720) using the same procedure and applied it on HUSK, DemGene and ADNI datasets.

#### (3) Performance evaluation of the PHS model

To evaluate the stratification performance of the developed model on the validation dataset and the independent test cohorts, we chose various metrics used in the literature. The Harrell’s Concordance index (C-index) metric is a generalization of the area under the ROC curve (AUC) that can take censored data into account in survival analysis [29]. We also examined the Hazard ratio (HR) between the risk groups stratified by PHS as defined elsewhere [17]. Furthermore, odds ratios (OR) of getting AD between risk groups until some ages (ranging from 65 to 85) were also examined. Specifically, we determined risk groups based on the PHS values of the samples and represented them as “high” whose PHS were in the top first 20 percentile (PHS>80%), “low” whose PHS is in the bottom 20 percentile (PHS<20%) and intermediate whose PHS is between (20%<PHS<80%). We also calculated corresponding hazard ratios (HR80/20) and odds ratios (OR80/20) between high and low-risk groups from age 65 to 85. In addition, we examined the risk groups with extremely high PHS values by comparing the top 2% and 5% percentiles to the bottom 50% and 20% (OR95/50, OR98/20 and OR95/20).

To report the confidence of the metrics on the EADB validation data, we ran the procedure defined here 100 times by randomly selecting train (80%) and validation data (20%), recorded the metrics for each run, and calculated the mean estimate of the metrics and corresponding standard deviations. To check the statistical significance of the performance increase compared to other models (such as Desikan Model) Two-Sample *t*-Test is used.

Additionally, we performed leave-one-country out analysis within EADB, iteratively training the model on data from all but one country and using the respective country for validation of model accuracy. After that, the entire EADB dataset was used to fit the ultimate model (EADB720) to test three independent out-sample cohorts: HUSK, DemGene and ADNI. Making use of bootstrapping 1000 times with replacement, we determined the corresponding standard deviation of the metrics for out-of-sample test datasets. The statistical significance of the results in out-sample cohorts are tested using bootstrapping for p-values as presented in [30].

## Results

We developed a new PHS model, termed EADB720, incorporating 720 genetic variants, including ε2 and ε4 using the Elastic Net-regularized Cox regression approach applied to EADB data. The performance of the model was evaluated on both validation data and independent test data. It is important to note that EADB720 model is obtained by using the whole EADB dataset and applied to out-sample cohorts. For internal EADB validation data, we performed a train and test procedure as defined before.

Initially for interval validation, we developed the PHS model on train data and calculated the corresponding metrics on the EADB validation data. Figure 1 shows Kaplan-Meier (KM) and Cox regression estimates for the EADB model on the EADB data. The figure demonstrates clear stratification among risk groups (log-rank test pval=2e-20). Specifically, the high-risk group (top 20 percent) reaches 0.5 AD-free survival probability at the age of 74 and the low-risk group (bottom 20 percent) reaches the same probability at 86 years.

**Figure 1.**
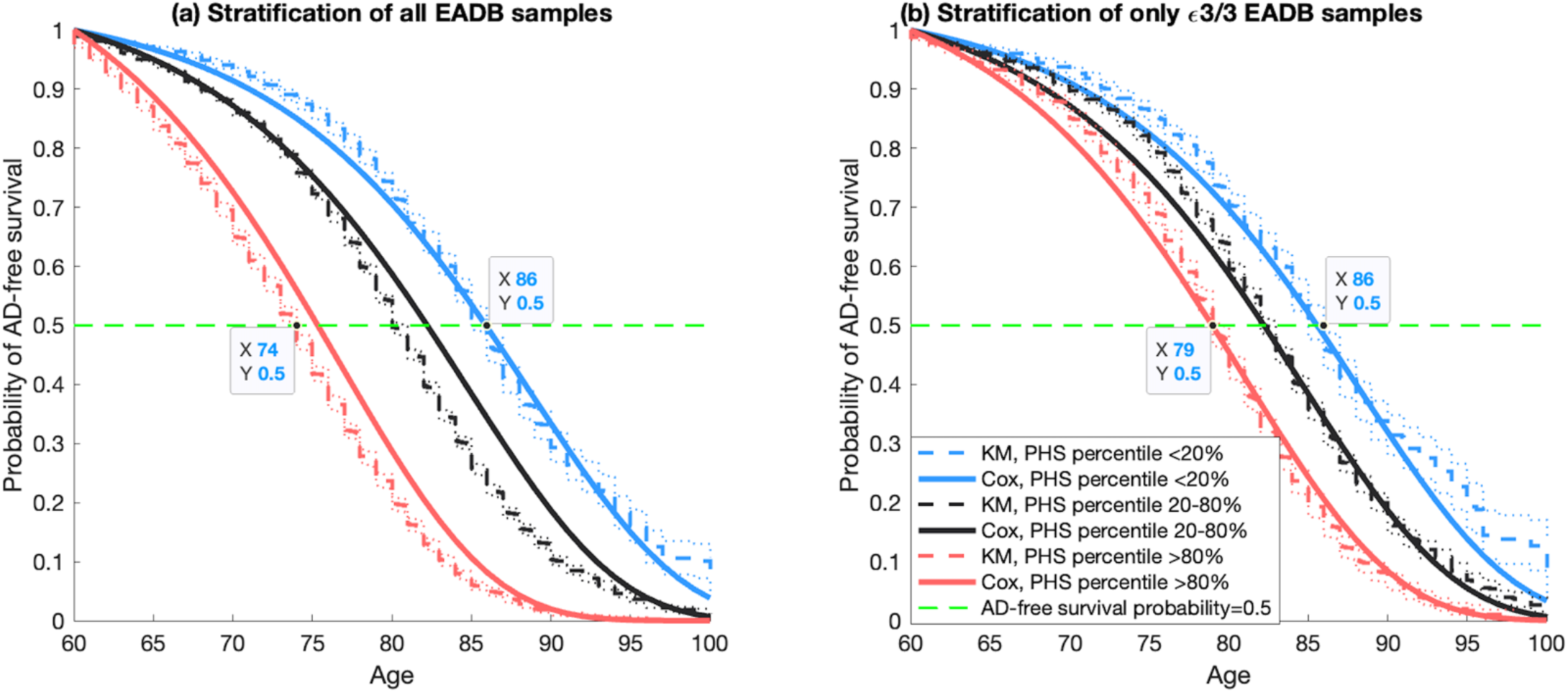
Kaplan-Meier (KM) curves and Cox proportional hazard model fits from the whole European Alzheimer & Dementia Biobank (EADB) dataset used to develop EADB Polygenic Hazard Score (PHS) model. a) Stratification of all EADB samples with respect to EADB720, b) Stratification of only *APOE* neutral (ε3/3) samples with respect to EADB720. The proportional hazard assumptions were checked based on graphical comparisons between KM estimates with 95% confidence intervals and Cox proportional hazard models (See Table 2 for numerical comparison). The vertical green-dashed line at 0.5 is to show the ages when risk groups reach 0.5 probability of Alzheimer’s disease (AD)-free survival.

**Table 2.** Comparison of our EADB model vs Desikan model with respect to C-index, Hazard Ratio (HR80/20), and Odds Ratio (OR80/20) using cross-validation within EADB samples. Odds ratios are shown across different age brackets. The numbers correspond to the mean estimates across 100 independent data splits into training and validation subsets. The numbers in the parenthesis are standard deviations. Comparisons are made for all validation samples, and in subsamples with neutral *APOE* status (ε3/3 samples).

| All validation samples | C-index | HR | OR<sub>80/20</sub> at 65 | OR<sub>80/20</sub> at 70 | OR<sub>80/20</sub> at 75 | OR<sub>80/20</sub> at 80 | OR<sub>80/20</sub> at 85 |
| --- | --- | --- | --- | --- | --- | --- | --- |
| EADB | 0.67<br>(0.005) | 2.63<br>(0.07) | 5.10<br>(1.26) | 6.55<br>(0.89) | 8.23<br>(0.88) | 9.13<br>(1.16) | 13.15<br>(2.10) |
| Desikan | 0.65<br>(0.005) | 2.17<br>(0.05) | 4.44<br>(0.98) | 5.49<br>(0.69) | 6.29<br>(0.69) | 6.71<br>(0.58) | 8.42<br>(1.12) |
| $\epsilon 3/3$ samples | | | | | | | |
| EADB | 0.57<br>(0.009) | 1.45<br>(0.06) | 1.95<br>(0.49) | 2.32<br>(0.40) | 2.77<br>(0.44) | 3.51<br>(0.48) | 3.52<br>(0.87) |
| Desikan | 0.54<br>(0.008) | 1.27<br>(0.04) | 1.50<br>(0.56) | 1.82<br>(0.48) | 1.87<br>(0.26) | 2.00<br>(0.27) | 2.04<br>(0.42) |

For validation data, we summarized the C-index, HR, and OR at ages from 65 to 85 in Table 2. Specifically, we examined the HR and OR between the high-risk and low-risk Groups, defined as the top and bottom 20% of PHS values (HR80/20 and OR80/20). Using the EADB model, we observed an HR80/20=2.63, and C-index=0.67(Table 2). The corresponding OR80/20 values are increasing from 5.10 to 13.15. We also examined the OR in higher percentiles of PHS. While OR80/20 varies from 5.10 and 13.15, the OR in the same age span is significantly higher for the samples with the highest percentiles such that OR95/20 increases from 11.32 to 40.25. The same metric for the samples with the highest two percentiles (OR98/20) increases from 14.65 to 52.91. More details on these groups can be found in Supplementary Table 1a-1c. Table 2 also presents a comparison between the EADB model with the Desikan PHS model using the same validation data. Our EADB model achieved statistically significant improvement with respect to C-index compared to Desikan model (0.67 vs. 0.65, two-Sample *t*-Test pval=1e-30) and improved OR80/20 values by up to 30 percent compared to the Desikan model. Similar trends are observed when only ε3/3 individuals are considered. In this case, the EADB720 model’s C-index is 0.57 vs. 0.54 for the Desikan model. Similarly, OR80/20 improved up to 50 percent compared to Desikan model.

While the *APOE* ε4 variant increases risk and may lead to earlier onset of AD, the *APOE* ε2 variant reduces AD risk or delays it. This effect can also be observed in Figure S4a, showing KM curves for EADB samples (whole cohort) stratified on *APOE* variants. These two variants’ effects with respect to the number of copies (0,1 or 2) on age of onset of AD were assumed to be linear in our model. However, a recent study suggested that homozygosity of ε4 represent a distinct genetic form of AD with a substantially higher risk [31]. We therefore checked whether the number of ε4 alleles affects the age of onset of AD linearly or not. To do this we calculated i) the OR of having two ε4 alleles (homozygote) to ε3/4 (having only one ε4), and ii) the OR of ε3/4 to ε3/3. These results were plotted from age 65 to 85 in Figure S4b. As shown, both ORs are almost steady from ages 65 to 85. This implies that the linearity assumption concerning the age of onset of AD and the number of ε4 alleles can be applicable in our dataset, and there is no need to use ε4 homozygosity as an additional predictor for age of onset.

While the *APOE* ε2 and ε4 variants are highly relevant for predicting the age of onset of AD, a significant proportion of the populations neither have ε2 nor ε4 variants (which corresponds to ε3/3 individuals). Therefore, for ε3/3 individuals, other genetic variants in a PHS model would become more important for differentiating between elevated or reduced risk of AD. We also examined the performance of our PHS model on only ε3/3 individuals. As shown in Figure 1b, the EADB720 model was able to stratify even among ε3/3 individuals. In ε3/3 individuals, the median AD-free survival time is 79 years for the top 20% PHS group, compared to 86 years for the bottom 20% PHS group. Therefore, even among ε3/3 individuals, PHS still captures AD onset and with 7 years difference with respect to 0.5 AD-free survival between the top 20 to bottom 20 percent risk groups. We further examined corresponding ORs at specific ages between risk groups to quantify the performance of the EADB720 model in ε3/3 individuals. As can be seen in Table 2, using EADB720, OR80/20 among ε3/3 individuals varies between 1.95 to 3.52 from the age of 65 to 85.

We also evaluated the performance of our approach using a leave-one-out (LOO) procedure based on the EADB dataset. By leaving out one of the countries in the EADB dataset, we iteratively performed training with the remaining countries, and used the excluded country samples for validation purposes (Table S3). Although performance differs by country due to possible case/control ratio, recruitment process differences etc., our model is able to stratify longitudinal AD risk of samples within countries. As suggested in [32], we also performed a sex-specific analysis by repeating the same procedure and developed a PHS models for men and women using a sex-matched dataset for training (Table S4). The performance of sex-specific models is similar with one another, with slightly better performance in the female-only cohort compared to the male cohort. On the other hand, compared to EADB model, there was not a significant improvement overall.

After performing the PHS analysis on the entire EADB dataset, we developed a PHS model which we applied and tested on out-of-sample datasets. In other words, we used the entire EADB dataset and repeated our training procedure presented in the “Statistical Analysis” section to test our PHS model on an independent dataset (DemGene, HUSK and ADNI) as presented in Table 3. Here, the EADB720 model outperformed the Desikan model in with respect to C-index in DemGene (pval=0.002), HUSK (pval=0.03) and ADNI (pval=0.01). There is up to 25 percent increase with respect to HR and 50 percent with respect to OR80/20 compared to the Desikan model. Validation with only ε3/3 individuals is more challenging and the performance among these individuals expectedly dropped. The EADB720 model however has a consistently superior performance compared to the Desikan model for ε3/3 individuals. Broader comparisons among these datasets are presented in Table S2c-S2e.

**Table 3.** PHS validation summary in DemGene, HUSK and ADNI datasets, showing C-index and Hazard Ratio (HR), and Odds Ratio (OR80/20) values with the EADB720 model applied on all and ε3/3 individuals. The Desikan model [15] performance is also included in the comparison. Broader results for these datasets are presented in (Table S2c-S2e).

| Dataset | Models | C-index<br>(for all<br>samples) | C-index<br>(for $\epsilon 3/3$<br>samples) | HR<br>(for all<br>samples) | HR<br>(for $\epsilon 3/3$<br>samples) | $OR_{80/20}$<br>at 80<br>(for all<br>samples) | $OR_{80/20}$<br>at 80<br>(for $\epsilon 3/3$<br>samples) |
| --- | --- | --- | --- | --- | --- | --- | --- |
| DemGene | EADB720 | 0.67<br>(0.006) | 0.55<br>(0.004) | 3.72<br>(0.31) | 1.38<br>(0.19) | 8.02<br>(0.84) | 1.62<br>(0.57) |
| DemGene | Desikan | 0.66<br>(0.006) | 0.52<br>(0.011) | 3.31<br>(0.23) | 1.22<br>(0.24) | 6.47<br>(0.99) | 1.12<br>(0.87) |
| HUSK | EADB720 | 0.72<br>(0.012) | 0.57<br>(0.012) | 5.20<br>(0.97) | 1.55<br>(0.94) | 7.53<br>(0.84) | 1.61<br>(0.64) |
| HUSK | Desikan | 0.70<br>(0.011) | 0.54<br>(0.012) | 4.30<br>(0.86) | 1.13<br>(0.91) | 5.66<br>(0.76) | 0.98<br>(0.82) |
| ADNI | EADB720 | 0.68<br>(0.013) | 0.59<br>(0.06) | 5.14<br>(0.57) | 1.90<br>(0.22) | 11.35<br>(1.02) | 2.88<br>(0.48) |
| ADNI | Desikan | 0.66<br>(0.015) | 0.57<br>(0.05) | 4.24<br>(0.49) | 1.63<br>(0.2) | 6.72<br>(0.98) | 1.77<br>(0.43) |

We also showed that EADB720 model outperforms AD Polygenic Risk Score (PRS) models which included the effect size obtained from the EADB GWAS [21] in Table S5. Such an improved accuracy was not observed for Desikan PHS model [33] . We also observed better accuracy compared with PRS from EADB PRS [21], PGC UKB [6] and meta-analysis obtained in this study and observed superior performance on independent datasets in Table S6.

## Discussion

We developed a new PHS model to predict age-associated risk of AD for European populations, using EADB data that includes samples from 15 different European countries. We used an Elastic Net-regularized Cox Proportional Hazards machine-learning framework that accounted for a total of 720 SNPs in the model (EADB720). We showed that the EADB720 model performed better than the previous PHS models and outperforms AD PRS models. Thus, the EADB720 PHS model has a greater potential to identify those at risk of AD and when they are at risk and thus increase the early diagnosis in AD.

We trained, validated and tested the EADB720 model in European cohort data and demonstrated how it outperforms the previous Desikan PHS model using several metrics (C-index, HR, OR). The performance was validated in EADB data, and then tested in independent European datasets, including Norwegian cohorts. The EADB720 PHS model obtained better accuracy than the previous Desikan PHS model and PRS models. We also developed sex-specific models using EADB data as done in [32], however, we did not observe any significant change in the overall performance. This can be either due to using lower sample size (roughly half of the total sample) and sex-independent metadata to pre-filter variants prior to PHS development. A follow-up study could consider developing a sex-specific model using sex-specific GWASes.

In both EADB720 and the Desikan model, the ε4 and ε2 variants have larger effect sizes than other variants. Recent findings [31] suggest that the homozygosity of ε4 is a distinct genetic form of AD. We therefore also examined the effect of ε4 on the age of onset and observed that the number of ε4 alleles has a somewhat linear effect on the age of onset. This suggests a dose-dependent effect of ε4 on age of onset, rather than a distinct genetic form of AD. While a majority of the population does not carry ε4 and/or ε2 variants, the PHS model should also be able to stratify the AD risk in the ε3/3 carrier group. Indeed, the EADB720 PHS model showed stratification among ε3/3 samples and outperformed the Desikan PHS model. Thus, the EADB720 model can also be used to provide useful decision support for drug treatment plans for ε3/3 patients, such as donanemab or lecanemab.

Improved genetic-based prediction for age of onset of AD may lead to early diagnosis to start treatment as well as decision support for initiating prevention at earlier ages. For example, amyloid beta accumulation in the brain is a hallmark of AD and can be detected before symptoms develop [8]. On the other hand, some of those who are amyloid positive, will never develop clinical symptoms of AD. Therefore, PHS might be an additional indicator to decide when (or whether) to start treatment in a preclinical phase based on amyloid positivity. Furthermore, using this PHS based prediction scheme can significantly help with better stratification of patients for clinical trials and optimizing the selection for existing FDA-approved drugs such as lecanemab and donanemab [11],[12]. This approach can also support the repurposing of existing drugs which may have neuroprotective effects as well focus on new drug development targeting specific pathways implicated in AD, such as amyloid-beta, tau, neuroinflammation and level of synaptic dysfunction [34].

Regularization based frameworks like LASSO regression have been used to identify SNPs for PRS of several disorders [35],[36]. It has also been used for PHS analysis with the Cox Proportional Hazard framework in prostate cancer and observed superior performance than previous stepwise selection approaches. LASSO provides more robust estimates than stepwise selection in cases with both a few large effects, as well as many small effects [18]. Therefore, we tested different regularization methods including LASSO, Ridge and ElasticNet and observed that the ElasticNet framework provides better prediction performance.

Our study has some limitations. Although our method has been validated and tested on different European datasets, it requires further validation in European and non-European cohorts. Furthermore, there is no consensus among the cohorts regarding the definition of AD onset and even within the cohort there are different variables which may be related to age of onset. We followed a similar procedure to pre-filter genome and phenome data in development and test cohorts, but their imputation pipelines and age distributions were not identical. These differences might lead to some variations on the results.

In summary, we developed a new PHS model (EADB720) that improved the accuracy of the prediction of age-associated AD risk in European samples. Our PHS model and framework may provide a clinical utility for early diagnosis and stratify intervention procedures in the early phase of AD.

## Data Availability

The study was approved by the institutional review board of each participating center and
written informed consent was obtained from each patient or a legal representative. EADB dataset is used
under the permission of REC# 2014/631. The HUSKment project was approved by REC-vest (permission
#2018/915). DemGene and ADNI datasets are used under the permission of REC# 2014/631. All other data All data produced in the present study are available upon reasonable request to the authors

## Acknowledgements

We thank all the participants in the studies. We also thank the high-performance computing service at the University of Lille. This work also used the TSD (Tjeneste for Sensitive Data) infrastructure, owned by the University of Oslo, operated and developed by the TSD service group at the University of Oslo, IT-Department (USIT), using resources provided by UNINETT Sigma2—the National Infrastructure for High Performance Computing and Data Storage in Norway (grant # NS9703S – p697). We want to acknowledge the Norwegian registry of persons assessed for cognitive symptoms (NorCog), patients and clinicians for providing access to patient data (and/or caregiver data, and/or biological material). This project is supported by Research Council of Norway (RCN) 324252, 344121 Research Council of Norway (#326813); EEA and Norway grant (#EEA-RO-NO-2018-0573) European Union’s Horizon 2020 Research and Innovation Programme (RealMent, Grant No. 964874) South-Eastern Norway Regional Health Authority (#2022073); Research Council of Norway (#324499) South-Eastern Norway Regional Health Authority (#2022073) Research Council of Norway (#223273, #248778), KG Jebsen Stiftelsen, The South-East Norway Regional Health Authority (#2022-087) Norwegian Health Association #22731 Research Council of Norway (RCN) #324499. Dr. Anders M. Dale is supported by the following grants from the National Institutes of Health (NIH): U24DA041123; R01AG076838; U24DA055330; and OT2HL161847.

## EADB

We thank the numerous participants, researchers, and staff from many studies who collected and contributed to the data. We thank the high performance computing service of the University of Lille. We thank all the CEA-CNRGH staff who contributed to sample preparation and genotyping for their excellent technical assistance. This research has been conducted using the UK Biobank Resource *under Application Number* 61054.

This work was supported by a grant (European Alzheimer DNA BioBank, EADB) from the EU Joint Programme – Neurodegenerative Disease Research (JPND). Inserm UMR1167 is also funded by Inserm, Institut Pasteur de Lille, the Lille Métropole Communauté Urbaine, the French government’s LABEX DISTALZ program (development of innovative strategies for a transdisciplinary approach to Alzheimer’s disease).

Additional support for EADB cohorts was provided by: The work for this manuscript was further supported by the CoSTREAM project (www.costream.eu) and funding from the European Union’s Horizon 2020 research and innovation programme under grant agreement No 667375. Italian Ministry of Health (Ricerca Corrente); Ministero dell’Istruzione, dell’Università e della Ricerca–MIUR project “Dipartimenti di Eccellenza 2018–2022” to Department of Neuroscience “Rita Levi Montalcini”, University of Torino (IR), and AIRAlzh Onlus-ANCC-COOP (SB); Partly supported by “Ministero della Salute”, I.R.C.C.S. Research Program, Ricerca Corrente 2018-2020, Linea n. 2 “Meccanismi genetici, predizione e terapie innovative delle malattie complesse” and by the “5 x 1000” voluntary contribution to the Fondazione I.R.C.C.S. Ospedale “Casa Sollievo della Sofferenza”; and RF-2018-12366665, Fondi per la ricerca 2019 (Sandro Sorbi). Copenhagen General Population Study (CGPS): We thank staff and participants of the CGPS for their important contributions. Karolinska Institutet AD cohort: Dr. Graff and co-authors of the Karolinska Institutet AD cohort report grants from Swedish Research Council (VR) 2015-02926, 2018-02754, 2015-06799, Swedish Alzheimer Foundation, Stockholm County Council ALF and resarch school, Karolinska Institutet StratNeuro, Swedish Demensfonden, and Swedish brain foundation, during the conduct of the study. ADGEN: This work was supported by Academy of Finland (grant numbers 307866); Sigrid Jusélius Foundation; the Strategic Neuroscience Funding of the University of Eastern Finland; EADB project in the JPNDCO-FUND program (grant number 301220). CBAS: Supported by the project no. LQ1605 from the National Program of Sustainability II (MEYS CR), Supported by Ministry of Health of the Czech Republic, grant nr. NV19-04-00270 (All rights reserved), Grant Agency of Charles University Grants No. 693018 and 654217; the Ministry of Health, Czech Republic―conceptual development of research organization, University Hospital Motol, Prague, Czech Republic Grant No. 00064203; the Czech Ministry of Health Project AZV Grant No. 16―27611A; and Institutional Support of Excellence 2. LF UK Grant No. 699012. CNRMAJ-Rouen: This study received fundings from the Centre National de Référence Malades Alzheimer Jeunes (CNRMAJ). The Finnish Geriatric Intervention Study for the Prevention of Cognitive Impairment and Disability (FINGER) data collection was supported by grants from the Academy of Finland, La Carita Foundation, Juho Vainio Foundation, Novo Nordisk Foundation, Finnish Social Insurance Institution, Ministry of Education and Culture Research Grants, Yrjö Jahnsson Foundation, Finnish Cultural Foundation South Osthrobothnia Regional Fund, and EVO/State Research Funding grants of University Hospitals of Kuopio, Oulu and Turku, Seinäjoki Central Hospital and Oulu City Hospital, Alzheimer’s Research & Prevention Foundation USA, AXA Research Fund, Knut and Alice Wallenberg Foundation Sweden, Center for Innovative Medicine (CIMED) at Karolinska Institutet Sweden, and Stiftelsen Stockholms sjukhem Sweden. FINGER cohort genotyping was funded by EADB project in the JPND CO-FUND (grant number 301220). Research at the Belgian EADB site is funded in part by the Alzheimer Research Foundation (SAO-FRA), The Research Foundation Flanders (FWO), and the University of Antwerp Research Fund. FK was supported by a postdoctoral fellowship (BOF 49758) from the University of Antwerp Research Fund and is a recipient of a postdoctoral fellowship from Brein Instituut. SNAC-K is financially supported by the Swedish Ministry of Health and Social Affairs, the participating County Councils and Municipalities, and the Swedish Research Council. BDR Bristol: We would like to thank the South West Dementia Brain Bank (SWDBB) for providing brain tissue for this study. The SWDBB is part of the Brains for Dementia Research programme, jointly funded by Alzheimer’s Research UK and Alzheimer’s Society and is supported by BRACE (Bristol Research into Alzheimer’s and Care of the Elderly) and the Medical Research Council. BDR Manchester: We would like to thank the Manchester Brain Bankfor providing brain tissue for this study. The Manchester Brain Bank is part of the Brains for Dementia Research programme, jointly funded by Alzheimer’s Research UK and Alzheimer’s Society. BDR KCL: Human post-mortem tissue was provided by the London Neurodegenerative Diseases Brain Bank which receives funding from the UK Medical Research Council and as part of the Brains for Dementia Research programme, jointly funded by Alzheimer’s Research UK and the Alzheimer’s Society. The CFAS Wales study was funded by the ESRC (RES-060-25-0060) and HEFCW as ‘Maintaining function and well-being in later life: a longitudinal cohort study’, (Principal Investigators: R.T Woods, L.Clare, G.Windle, V. Burholt, J. Philips, C. Brayne, C. McCracken, K. Bennett, F. Matthews). We are grateful to the NISCHR Clinical Research Centre for their assistance in tracing participants and in interviewing and in collecting blood samples, and to general practices in the study areas for their cooperation. MRC: We thank all individuals who participated in this study. Cardiff University was supported by the Alzheimer’s Society (AS; grant RF014/164) and the Medical Research Council (MRC; grants G0801418/1, MR/K013041/1, MR/L023784/1) (R. Sims is an AS Research Fellow). Cardiff University was also supported by the European Joint Programme for Neurodegenerative Disease (JPND; grant MR/L501517/1), Alzheimer’s Research UK (ARUK; grant ARUK-PG2014-1), the Welsh Assembly Government (grant SGR544:CADR), Brain’s for dementia Research and a donation from the Moondance Charitable Foundation. Cardiff University acknowledges the support of the UK Dementia Research Institute, of which J. Williams is an associate director. Cambridge University acknowledges support from the MRC. Patient recruitment for the MRC Prion Unit/UCL Department of Neurodegenerative Disease collection was supported by the UCLH/UCL Biomedical Centre and NIHR Queen Square Dementia Biomedical Research Unit. The University of Southampton acknowledges support from the AS. King’s College London was supported by the NIHR Biomedical Research Centre for Mental Health and the Biomedical Research Unit for Dementia at the South London and Maudsley NHS Foundation Trust and by King’s College London and the MRC. ARUK and the Big Lottery Fund provided support to Nottingham University. Alfredo Ramirez: Part of the work was funded by the JPND EADB grant (German Federal Ministry of Education and Research (BMBF) grant: 01ED1619A). German Study on Ageing, Cognition and Dementia in Primary Care Patients (AgeCoDe): This study/publication is part of the German Research Network on Dementia (KND), the German Research Network on Degenerative Dementia (KNDD; German Study on Ageing, Cognition and Dementia in Primary Care Patients; AgeCoDe), and the Health Service Research Initiative (Study on Needs, health service use, costs and health-related quality of life in a large sample of oldestold primary care patients (85+; AgeQualiDe)) and was funded by the German Federal Ministry of Education and Research (grants KND: 01GI0102, 01GI0420, 01GI0422, 01GI0423, 01GI0429, 01GI0431, 01GI0433, 01GI0434; grants KNDD: 01GI0710, 01GI0711, 01GI0712, 01GI0713, 01GI0714, 01GI0715, 01GI0716; grants Health Service Research Initiative: 01GY1322A, 01GY1322B, 01GY1322C, 01GY1322D, 01GY1322E, 01GY1322F, 01GY1322G). In addition, this publication was supported by the German Federal Ministry of Education and Research (Grant numbers: PreADAPT project 01ED2007A and DESCARTES project 01EK2102B to Alfredo Ramirez, 01EK2102A to Anja Schneider, and 01EK2102C to Michael Wagner. VITA study: The support of the Ludwig Boltzmann Society and the AFI Germany have supported the VITA study. The former VITA study group should be acknowledged: W. Danielczyk, G. Gatterer, K Jellinger, S Jugwirth, KH Tragl, S Zehetmayer. Vogel Study: This work was financed by a research grant of the ‘‘Vogelstiftung Dr. Eckernkamp’’. HELIAD study: This study was supported by the grants: IIRG-09-133014 from the Alzheimer’s Association, 189 10276/8/9/2011 from the ESPA-EU program Excellence Grant (ARISTEIA) and the ΔΥ2β/οικ.51657/14.4.2009 of the Ministry for Health and Social Solidarity (Greece). Biobank Department of Psychiatry, UMG: Prof. Jens Wiltfang is supported by an Ilídio Pinho professorship and iBiMED (UID/BIM/04501/2013), and FCT project PTDC/DTP_PIC/5587/2014 at the University of Aveiro, Portugal. Lausanne study: This work was supported by grants from the Swiss National Research Foundation (SNF 320030_141179). PAGES study: Harald Hampel is an employee of Eisai Inc. During part of this work he was supported by the AXA Research Fund, the “Fondation partenariale Sorbonne Université” and the “Fondation pour la Recherche sur Alzheimer”, Paris, France. Mannheim, Germany Biobank: Department of geriatric Psychiatry, Central Institute for Mental Health, Mannheim, University of Heidelberg, Germany. Genotyping for the Swedish Twin Studies of Aging was supported by NIH/NIA grant R01 AG037985. Genotyping in TwinGene was supported by NIH/NIDDK U01 DK066134. WvdF is recipient of Joint Programming for Neurodegenerative Diseases (JPND) grants PERADES (ANR-13-JPRF-0001) and EADB (733051061). Gothenburg Birth Cohort (GBC) Studies: We would like to thank UCL Genomics for performing the genotyping analyses. The studies were supported by The Stena Foundation, The Swedish Research Council (2015-02830, 2013-8717), The Swedish Research Council for Health, Working Life and Wellfare (2013-1202, 2005-0762, 2008-1210, 2013-2300, 2013-2496, 2013-0475), The Brain Foundation, Sahlgrenska University Hospital (ALF), The Alzheimer’s Association (IIRG-03-6168), The Alzheimer’s Association Zenith Award (ZEN-01-3151), Eivind och Elsa K:son Sylvans Stiftelse, The Swedish Alzheimer Foundation. Clinical AD, Sweden: We would like to thank UCL Genomics for performing the genotyping analyses. Barcelona Brain Biobank: Brain Donors of the Neurological Tissue Bank of the Biobanc-Hospital Clinic-IDIBAPS and their families for their generosity. Hospital Clínic de Barcelona Spanish Ministry of Economy and Competitiveness-Instituto de Salud Carlos III and Fondo Europeo de Desarrollo Regional (FEDER), Unión Europea, “Una manera de hacer Europa” grants (PI16/0235 to Dr. R. Sánchez-Valle and PI17/00670 to Dr. A.Antonelli). AA is funded by Departament de Salut de la Generalitat de Catalunya, PERIS 2016-2020 (SLT002/16/00329). Work at JP-T laboratory was possible thanks to funding from Ciberned and generous gifts from Consuelo Cervera Yuste and Juan Manuel Moreno Cervera. Sydney Memory and Ageing Study (Sydney MAS): We gratefully acknowledge and thank the following for their contributions to Sydney MAS: participants, their supporters and the Sydney MAS Research Team (current and former staff and students). Funding was awarded from the Australian National Health and Medical Research Council (NHMRC) Program Grants (350833, 568969, 109308). AddNeuroMed consortium was led by Simon Lovestone, Bruno Vellas, Patrizia Mecocci, Magda Tsolaki, Iwona Kłoszewska, Hilkka Soininen. This work was supported by InnoMed (Innovative Medicines in Europe), an integrated project funded by the European Union of the Sixth Framework program priority (FP6-2004-LIFESCIHEALTH-5). Oviedo: This work was partly supported by Grant from Fondo de Investigaciones Sanitarias-Fondos FEDER EuropeanUnion to Victoria Alvarez PI15/00878. Pascual Sánchez-Juan is supported by CIBERNED and Carlos III Institute of Health, Spain (PI08/0139, PI12/02288, and PI16/01652), jointly funded by Fondo Europeo de Desarrollo Regional (FEDER), Unión Europea, “Una manera de hacer Europa”. Project MinE: The ProjectMinE study was supported by the ALS Foundation Netherlands and the MND association (UK) (Project MinE, www.projectmine.com). The SPIN cohort: We are indebted to patients and their families for their participation in the “Sant Pau Initiative on Neurodegeneration cohort”, at the Sant Pau Hospital (Barcelona). This is a multimodal research cohort for biomarker discovery and validation that is partially funded by Generalitat de Catalunya (2017 SGR 547 to JC), as well as from the Institute of Health Carlos III-Subdirección General de Evaluación and the Fondo Europeo de Desarrollo Regional (FEDER-“Una manera de Hacer Europa”) (grants PI11/02526, PI14/01126, and PI17/01019 to JF; PI17/01895 to AL), and the Centro de Investigación Biomédica en Red Enfermedades Neurodegenerativas programme (Program 1, Alzheimer Disease to AL). We would also like to thank the Fundació Bancària Obra Social La Caixa (DABNI project) to JF and AL; and Fundación BBVA (to AL), for their support in funding this follow-up study. Adolfo López de Munain is supported by Fundación Salud 2000 (PI2013156), CIBERNED and Diputación Foral de Gipuzkoa (Exp.114/17). This study was supported by the Italian Ministry of Health (Ricerca Corrente) (RG, BB). This work was supported by grants from the Swiss National Science Foundation (grant number 320030_141179; 320030_204886) and from the Synapsis Foundation - Dementia Research Switzerland (grant number 2017-PI01) to J. Popp. The Bulgarian cohort was partially supported by the European Union-NextGenerationEU (National Recovery and Resilience Plan of the Republic of Bulgaria, grant number BG-RRP-2.004-0004-C01), and the Bulgarian National Science Fund (BNSF, KP-06-N53/3 and DN 03/12).

## Gra@ce

The Genome Research @ Fundació ACE project (GR@ACE) is supported by Grifols SA, Fundación bancaria ‘La Caixa’, Fundació ACE, and CIBERNED (Centro de Investigación Biomédica en Red Enfermedades Neurodegenerativas (Program 1, Alzheimer Disease to MB and AR)). A.R. and M.B. receive support from the European Union/EFPIA Innovative Medicines Initiative Joint undertaking ADAPTED and MOPEAD projects (grant numbers 115975 and 115985, respectively). M.B. and A.R. are also supported by national grants PI13/02434, PI16/01861, PI17/01474 and PI19/01240. Acción Estratégica en Salud is integrated into the Spanish National R + D + I Plan and funded by ISCIII (Instituto de Salud Carlos III)–Subdirección General de Evaluación and the Fondo Europeo de Desarrollo Regional (FEDER–‘Una manera de hacer Europa’). Some control samples and data from patients included in this study were provided in part by the National DNA Bank Carlos III (www.bancoadn.org, University of Salamanca, Spain) and Hospital Universitario Virgen de Valme (Sevilla, Spain); they were processed following standard operating procedures with the appropriate approval of the Ethical and Scientific Committee. The present work has been performed as part of the doctoral program of I. de Rojas at the Universitat de Barcelona (Barcelona, Spain).

## EADI

This work has been developed and supported by the LABEX (laboratory of excellence program investment for the future) DISTALZ grant (Development of Innovative Strategies for a Transdisciplinary approach to ALZheimer’s disease) including funding from MEL (Metropole européenne de Lille), ERDF (European Regional Development Fund) and Conseil Régional Nord Pas de Calais. This work was supported by INSERM, the National Foundation for Alzheimer’s disease and related disorders, the Institut Pasteur de Lille and the Centre National de Recherche en Génomique Humaine, CEA, the JPND PERADES, the Laboratory of Excellence GENMED (Medical Genomics) grant no. ANR-10-LABX-0013 managed by the National Research Agency (ANR) part of the Investment for the Future program, and the FP7 AgedBrainSysBio. The Three-City Study was performed as part of collaboration between the Institut National de la Santé et de la Recherche Médicale (Inserm), the Victor Segalen Bordeaux II University and Sanofi-Synthélabo. The Fondation pour la Recherche Médicale funded the preparation and initiation of the study. The 3C Study was also funded by the Caisse Nationale Maladie des Travailleurs Salariés, Direction Générale de la Santé, MGEN, Institut de la Longévité, Agence Française de Sécurité Sanitaire des Produits de Santé, the Aquitaine and Bourgogne Regional Councils, Agence Nationale de la Recherche, ANR supported the COGINUT and COVADIS projects. Fondation de France and the joint French Ministry of Research/INSERM “Cohortes et collections de données biologiques” programme. Lille Génopôle received an unconditional grant from Eisai. The Three-city biological bank was developed and maintained by the laboratory for genomic analysis LAG-BRC - Institut Pasteur de Lille.

## GERAD

We thank all individuals who participated in this study. Cardiff University was supported by the Wellcome Trust, Alzheimer’s Society (AS; grant RF014/164), the Medical Research Council (MRC; grants G0801418/1, MR/K013041/1, MR/L023784/1), the European Joint Programme for Neurodegenerative Disease (JPND, grant MR/L501517/1), Alzheimer’s Research UK (ARUK, grant ARUK-PG2014-1), Welsh Assembly Government (grant SGR544:CADR), a donation from the Moondance Charitable Foundation, UK Dementia’s Platform (DPUK, reference MR/L023784/1), and the UK Dementia Research Institute at Cardiff. Cambridge University acknowledges support from the MRC. ARUK supported sample collections at the Kings College London, the South West Dementia Bank, Universities of Cambridge, Nottingham, Manchester and Belfast. King’s College London was supported by the NIHR Biomedical Research Centre for Mental Health and Biomedical Research Unit for Dementia at the South London and Maudsley NHS Foundation Trust and Kings College London and the MRC. Alzheimer’s Research UK (ARUK) and the Big Lottery Fund provided support to Nottingham University. Ulster Garden Villages, AS, ARUK, American Federation for Aging Research, NI R&D Office and the Royal College of Physicians/Dunhill Medical Trust provided support for Queen’s University, Belfast. The University of Southampton acknowledges support from the AS. The MRC and Mercer’s Institute for Research on Ageing supported the Trinity College group. DCR is a Wellcome Trust Principal Research fellow. The South West Dementia Brain Bank acknowledges support from Bristol Research into Alzheimer’s and Care of the Elderly. The Charles Wolfson Charitable Trust supported the OPTIMA group. Washington University was funded by NIH grants, Barnes Jewish Foundation and the Charles and Joanne Knight Alzheimer’s Research Initiative. Patient recruitment for the MRC Prion Unit/UCL Department of Neurodegenerative Disease collection was supported by the UCLH/UCL Biomedical Research Centre and their work was supported by the NIHR Queen Square Dementia BRU, the Alzheimer’s Research UK and the Alzheimer’s Society. LASER-AD was funded by Lundbeck SA. The AgeCoDe study group was supported by the German Federal Ministry for Education and Research grants 01 GI 0710, 01 GI 0712, 01 GI 0713, 01 GI 0714, 01 GI 0715, 01 GI 0716, 01 GI 0717. Genotyping of the Bonn case-control sample was funded by the German centre for Neurodegenerative Diseases (DZNE), Germany. The GERAD Consortium also used samples ascertained by the NIMH AD Genetics Initiative. HH was supported by a grant of the Katharina-Hardt-Foundation, Bad Homburg vor der Höhe, Germany. The KORA F4 studies were financed by Helmholtz Zentrum München; German Research Center for Environmental Health; BMBF; German National Genome Research Network and the Munich Center of Health Sciences. The Heinz Nixdorf Recall cohort was funded by the Heinz Nixdorf Foundation (Dr. Jur. G.Schmidt, Chairman) and BMBF. We acknowledge use of genotype data from the 1958 Birth Cohort collection and National Blood Service, funded by the MRC and the Wellcome Trust which was genotyped by the Wellcome Trust Case Control Consortium and the Type-1 Diabetes Genetics Consortium, sponsored by the National Institute of Diabetes and Digestive and Kidney Diseases, National Institute of Allergy and Infectious Diseases, National Human Genome Research Institute, National Institute of Child Health and Human Development and Juvenile Diabetes Research Foundation International. The project is also supported through the following funding organisations under the aegis of JPND - www.jpnd.eu (United Kingdom, Medical Research Council (MR/L501529/1; MR/R024804/1) and Economic and Social Research Council (ES/L008238/1)) and through the Motor Neurone Disease Association. This study represents independent research part funded by the National Institute for Health Research (NIHR) Biomedical Research Centre at South London and Maudsley NHS Foundation Trust and King’s College London. Prof Jens Wiltfang is supported by an Ilídio Pinho professorship and iBiMED (UID/BIM/04501/2013), at the University of Aveiro, Portugal.

### Rotterdam study

Rotterdam (RS). This study was funded by the Netherlands Organisation for Health Research and Development (ZonMW) as part of the Joint Programming for Neurological Disease (JPND)as part of the PERADES Program (Defining Genetic Polygenic, and Environmental Risk for Alzheimer’s disease using multiple powerful cohorts, focused Epigenetics and Stem cell metabolomics), Project number 733051021. This work was funded also by the European Union Innovative Medicine Initiative (IMI) programme under grant agreement No. 115975 as part of the Alzheimer’s Disease Apolipoprotein Pathology for Treatment Elucidation and Development (ADAPTED, https://www.imi-adapted.eu);and the European Union’s Horizon 2020 research and innovation programme as part of the Common mechanisms and pathways in Stroke and Alzheimer’s disease CoSTREAM project (www.costream.eu, grant agreement No. 667375). The current study is supported by the Deltaplan Dementie and Memorabel supported by ZonMW (Project number 733050814) and Alzheimer Nederland. The Rotterdam Study is funded by Erasmus Medical Center and Erasmus University, Rotterdam, Netherlands Organization for the Health Research and Development (ZonMw), the Research Institute for Diseases in the Elderly (RIDE), the Ministry of Education, Culture and Science, the Ministry for Health, Welfare and Sports, the European Commission (DG XII), and the Municipality of Rotterdam. The authors are grateful to the study participants, the staff from the Rotterdam Study and the participating general practitioners and pharmacists. The generation and management of GWAS genotype data for the Rotterdam Study (RS-I, RS-II, RS-III) was executed by the Human Genotyping Facility of the Genetic Laboratory of the Department of Internal Medicine, Erasmus MC, Rotterdam, The Netherlands. The GWAS datasets are supported by the Netherlands Organization of Scientific Research NWO Investments (Project number 175.010.2005.011, 911-03-012), the Genetic Laboratory of the Department of Internal Medicine, Erasmus MC, the Research Institute for Diseases in the Elderly (014-93-015; RIDE2), the Netherlands Genomics Initiative (NGI)/Netherlands Organization for Scientific Research (NWO) Netherlands Consortium for Healthy Aging (NCHA), project number 050-060-810. We thank Pascal Arp, Mila Jhamai, Marijn Verkerk, Lizbeth Herrera and Marjolein Peters, MSc, and Carolina Medina-Gomez, MSc, for their help in creating the GWAS database, and Karol Estrada, PhD, Yurii Aulchenko, PhD, and Carolina Medina-Gomez, MSc, for the creation and analysis of imputed data.

### DemGene

The project has received funding from The Research Council of Norway (RCN) Grant Nos. 213837, 223273, 225989, 248778, and 251134 and EU JPND Program ApGeM RCN Grant No. 237250, the South-East Norway Health Authority Grant No. 2013-123, the Norwegian Health Association, and KG Jebsen Foundation. The RCN FRIPRO Mobility grant scheme (FRICON) is co-funded by the European Union’s Seventh Framework Programme for research, technological development and demonstration under Marie Curie grant agreement No 608695. European Community’s grant PIAPP-GA-2011-286213 PsychDPC.

### Bonn study

This group would like to thank Dr. Heike Koelsch for her scientific support. The Bonn group was funded by the German Federal Ministry of Education and Research (BMBF): Competence Network Dementia (CND) grant number 01GI0102, 01GI0711, 01GI042

### Data statement

The study was approved by the institutional review board of each participating center and written informed consent was obtained from each patient or a legal representative. EADB dataset is used under the permission of REC# 2014/631. The HUSKment project was approved by REC-vest (permission #2018/915). DemGene and ADNI datasets are used under the permission of REC# 2014/631. EADB720 PHS model can be accesible via this <u>link</u>.

## Author contributions

Cohort investigator: J.P.

Cohort PI: A.C., A.D., A.D.S., A.G., A.L., A.P.P., A.R., A.Ram., A.S., A.Sp., B.B., B.K., B.W., C.B., C.F., C.H., C.L., D.R., D.W., E.G., E.R.R., G.B., G.G.R., G.W., H.S., I.R., J.D., J.H., J.Ha., J.C.L., J.K., J.P.T., J.v.S., J.W., K.J., K.S., L.F., L.G., L.H., L.M.R., L.T., L.Tr., M.B., M.C., M.D., M.M.G., M.R., M.S., M.Sc., M.T., N.F.F., N.S., P.B., P.D.L.G., P.F., P.F.R., P.M., P.P., R.F.S., R.G., R.V., S.E., S.M., S.Me., S.R.H., S.T., T.F., V.F., Y.C., Y.P., L.O.W.

Data Analysis: A.C., A.R., B.C.A., E.H., G.N., I.J.B., K.P., O.F., O.S.G., R.M., S.B., S.H., S.D., Y.C.

Data Curation: A.C., A.E., B.C.A., B.W., C.B., D.S., E.G., F.K., F.R., G.O., G.R.G., I.d.R., J.F., K.P., L.G., L.O.W., M.A., M.M.B., M.R.R., M.Y., O.B., P.F., R.P., S.B., S.V.B., T.d.S., V.F., Y.C.

Data Preparation: A.C.A., A.C., A.Ca., A.R., A.S., B., C.B., C.L., E.A., E.G., E.H., F.K., F.P.P., F.R., G.N., G.S., H.W., I.A., I.d.R., I.T.M., J.D., J.M., J.W., K.P., K.S., L.A., L.A.S., L.F., L.G., L.H., L.K., M.E., M.E.S., M.H., M.S., N.B., N.T.F., P.G.G., P.H., R.H., R.S., R.V., S.B.S., S.E., S.H.H., S.L.G., S.T., T.T.F., V.A., V.G.

Genetics analysis: B.C.A.

Genetics collection: A.A., A.B.P., A.C., A.C.C., A.D., A.L., A.LO.D.M., A.M., A.O., A.R., A.Ram., A.S., B.M., B.W., C.B., C.C., C.F., C.L., C.P., D.G., D.R., D.S., D.W., E.C., E.F.M., E.G., E.R.R., E.Ru., F.C.G., F.I., F.P., G.N., H.W., I.d.R., I.Q., J.H., J.M., J.M.G., J.S., J.W., K.J.B., K.N., K.Y.M., L.A., L.M.R., L.M., L.T., M.B.S.A., M.C., M.E.S., M.G.V., M.H., M.M., M.M.N., M.O., M.S., N.S., P.A.P., P.C., P.G.K., P.M.L., P.T., R.H., S.B., S.D., S.H.H., S.J., S.M., S.Me., T.P., V.A., V.F., V.G., V.N.

Manuscript revision/editing: A.C., A.C.G., A.D., A.L., A.P.P., A.R., A.Ram., A.S., B., B.B., B.K., B.N., C.B., C.L., C.P., D.A., D.R., E.H., E.R.R., F.K., F.P., F.R., G.B., G.C.P., G.N., H.S., I.B., I.J.B., I.S., J.D., J.F., J.H., J.Ha., J.C.L., J.M.S., J.Q.T., J.S., J.v.S., J.W., K.J.B., K.N., K.S., L.F., L.G., L.H., L.K., L.M.R., L.T., M.A., M.B., M.C., M.D., M.E., M.H., M.M., M.R.R., M.S., M.Sc., M.T., M.V., M.W., N.B., N.S., N.Sc., O.A., O.B., O.F., O.S.G., P.A., P.B., P.F., P.F.R., P.G.K., P.H., P.M., P.R., P.S.J., R.F.S., R.G., R.H., R.P., S.B., S.D., S.F., S.H., S.M., T.d.S., T.F., T.T.F., V.A.

Methodology: A.C., A.R., B.C.A., C.C.P., E.H., I.J.B., J.F., L.O.W., M.A., M.R.R., O.F., O.K.D., O.M., O.S.G., P.A., S.B., S.D., T.T.F., An.,D., O.A.A.

Phenotypic Data Collection: A.A., A.B.P., A.D., A.H., A.L., A.LO.D.M., A.M.H., A.O., A.P.P., A.R., A.S., B.B., B.N., B.W., C.B., C.H., C.L., C.P., D.R., D.S., D.W., E.A., E.F.M., E.G., E.G.R., E.R.R., E.Ru., F.M., F.P., F.R., F.S., F.T., G.G., G.H., G.R.G., G.S., H.S., I.A., I.R., J.C.P., J.D., J.M.G., J.M.M., J.P., J.S., J.W., K.H., K.Y.M., L.A., L.H., L.K., L.M.D., L.P., L.T., M.B.S.A., M.D., M.G.V., M.H., M.M., M.M.G., M.O., M.P., M.R., M.S., N.B., N.F., N.S., N.Sc., O.G., P.A., P.B., P.C., P.F., P.G.K., P.M., P.P., R.F.S., R.G., R.H., R.P., R.V., S.B., S.C., S.E., S.F., S.H., S.M., S.Me., S.P., V.G.

Supervision

A.C., A.R., C.F., G.B., J.P., L.D.T., L.G., M.I., M.T., O.F., P.A.P., P.F.R., P.G.K., P.M., An.,D., O.A.A.

Writing team

B.C.A., O.F., S.B.

## Supplementary Notes

QC and imputation HUSK

HUSK participants were genotyped at deCODE Genetics (https://www.decode.com/) in two batches (10,083 and 25,432 individuals in batch 1 and 2 respectively) using customized Illumina GSA v3 array (687316 genotyped variants). Prior to phasing and imputation genotyping data were QCed separately for each batch using plink2 [20]. QC steps for variants included removal of indels, non-ATGC, strand ambiguous, poorly genotyped (plink2’s option: --geno 0.05), non-autosomal and rare (--maf 0.05) variants as well as variants with extreme deviation from Hardy-Weinberg equilibrium (--hwe 1E-50). Chromosomal strand and allele order for the remaining variants were aligned based on the HGDP + 1KG reference panel from gnomAD (https://gnomad.broadinstitute.org/downloads#v3-hgdp-1kg) and variants with alleles which were not possible to align to the reference as well as variants with MAF deviating from the reference (HGDP + 1KG) by more than 30% were removed. Resulting genotypes from two batches were merged together, variants with high missingness (--geno 0.02) and individuals with poor genotyping rate (--mind 0.05) were removed from the merged data. Combined dataset was then phased and imputed with Beagle 5.4 (version 27May24.118) using the HGDP + 1KG reference panel. Imputed data were further QCed by removing variants with low imputation quality (Beagle’s DR2<0.5) and variants with minor allele frequency below 0.5% (--maf 0.005). The resulting dataset contained 10,531,075 variants and 35,476 individuals.

### DemGene

Genotype data were phased and imputed using Beagle 5.5, version 17Dec24.224 with HGDP + 1KG reference panel [37]. Prior to phasing and imputation each batch was QCed separately. Briefly, using plink2 we excluded variants with high rate of missing genotype calls (--geno 0.05), variants with extreme deviation from Hardy-Weinberg equilibrium (--hwe 1E-50) and individuals with high genotyping missingness (--mind 0.2), then we applied the script from MxCarthy Group v4.3 (https://www.chg.ox.ac.uk/~wrayner/tools/) which aligns the strand, position, and ref/alt assignments according to the TOPMed imputation panel, and removes strand ambiguous variants and variants which cannot be aligned on the reference panel. Per-batch imputed vcf files obtained from the TOPMed server were merged using bcftools. Merged dataset was further QCed by removing variants with low imputation quality (Minimac’s R2<0.8), high rate of missing genotype calls (--geno 0.02), extreme deviation from Hardy-Weinberg equilibrium (--hwe 1E-50), low minor allele frequency (--mac 1) and individuals with high genotyping missingness (--mind 0.2). The resulting dataset contained 7,565,967 variants and 38,220 individuals.

### ADNI

To ensure the integrity and reliability of the genotype data used in this study, we performed a comprehensive QC process on datasets obtained from the Alzheimer’s Disease Neuroimaging Initiative (ADNI). The datasets included five batches. The QC steps were designed to filter out samples and SNPs with low-quality genotype data, and harmonize the data across batches. Prior to QC, ADNI1 batch was lifted over from hg18 to hg19. Phasing and imputation were carried out using the TOPMed Imputation Server, which employs Eagle for phasing and Minimac for imputation. The reference panel was set to ’All,’ and the R² threshold was set to 0.3. After imputation, imputed VCF files from each batch were merged, and the minimum R² score for each variant was retained across all batches in the merged files. The merged chromosome files were then converted to PLINK format, and all chromosomes were subsequently combined. The final dataset comprises 11,377,441 variants and 2,039 samples.

## Supplementary Material

**Fig S1.**
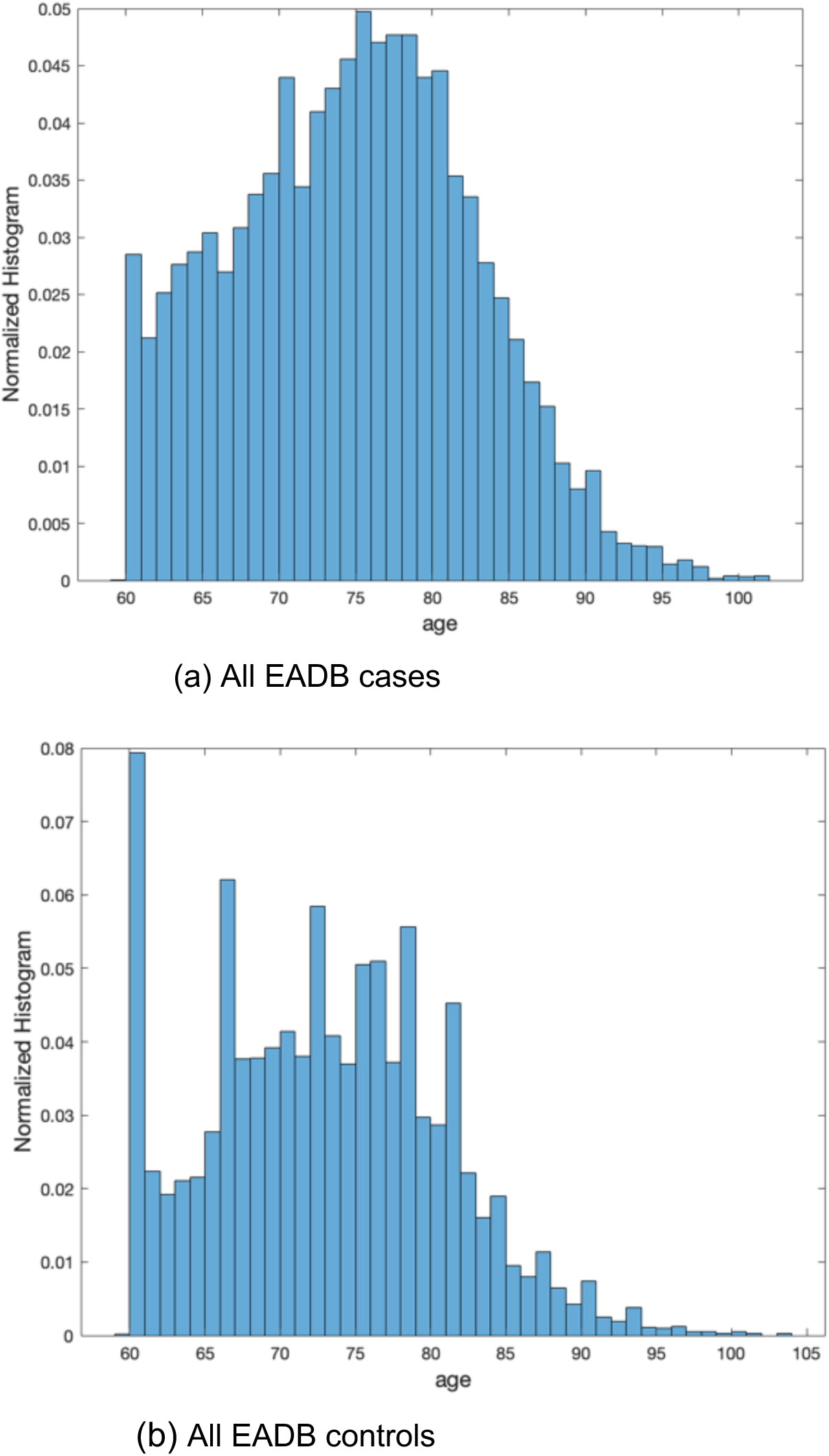
Age of onset of AD distribution for cases and age of last follow up distribution for controls for EADB dataset after filtering mentioned in the manuscript. For cases, “age_at_onset” variable is used to determine age of onset. For controls, “age_last_exam” variable is used for age of last examination. If this variable is not available, then “age_baseline” variable is used instead.

**Fig S2.**
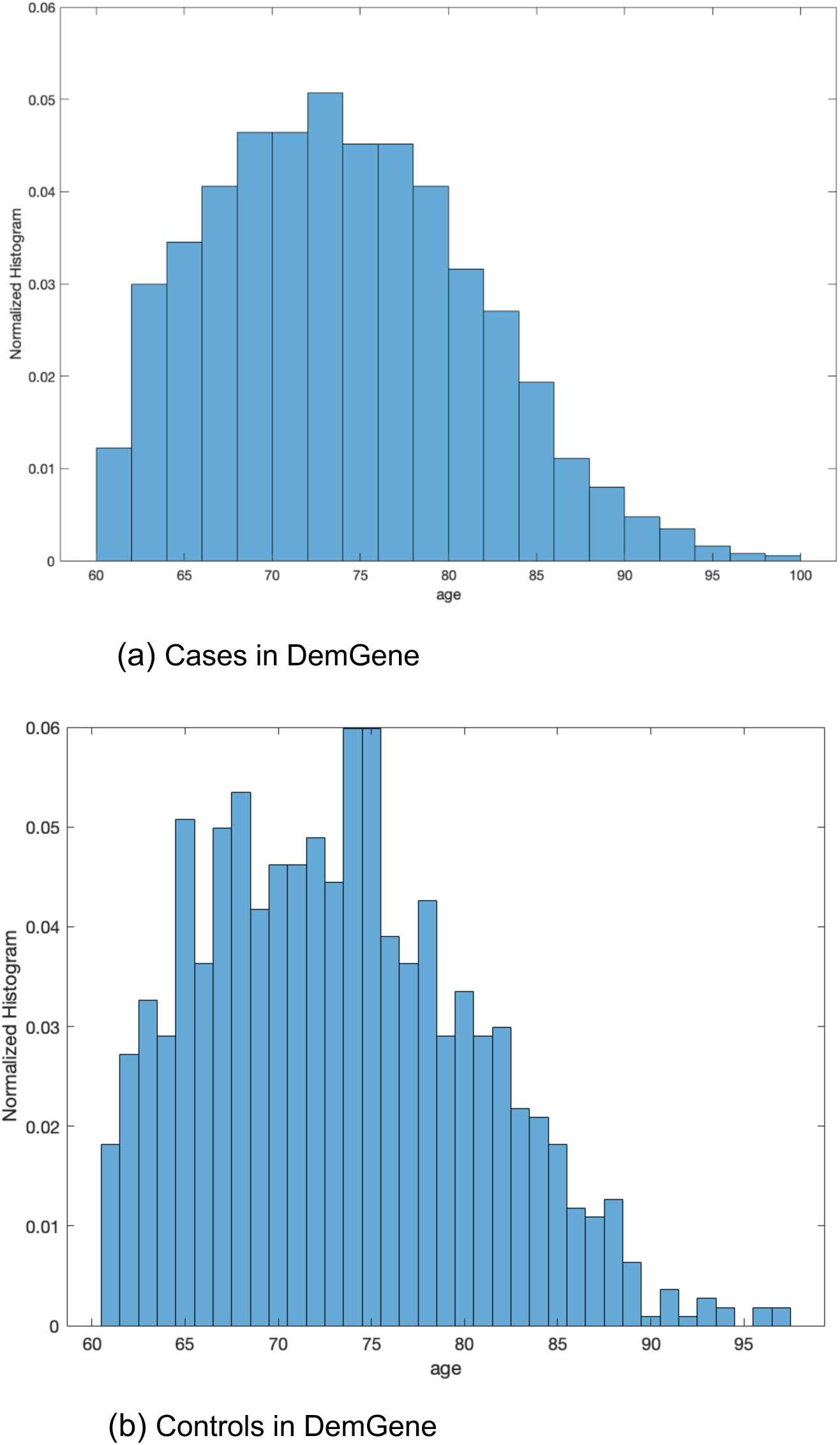
Age of onset of AD distribution for cases and age of last follow up distribution for controls for Demgene data. In DemGene dataset, there are 3 different variables related to age of onset: i) “demo_age_diagnosis”, ii) “demo_age_onset”, iii) “demo_age_symptom_start”. Since for some case samples, some of these variables were missing, we were using these variables with their priority order as listed here. For controls, we used “demo_age” variable for age of last follow-up.

**Fig S3.**
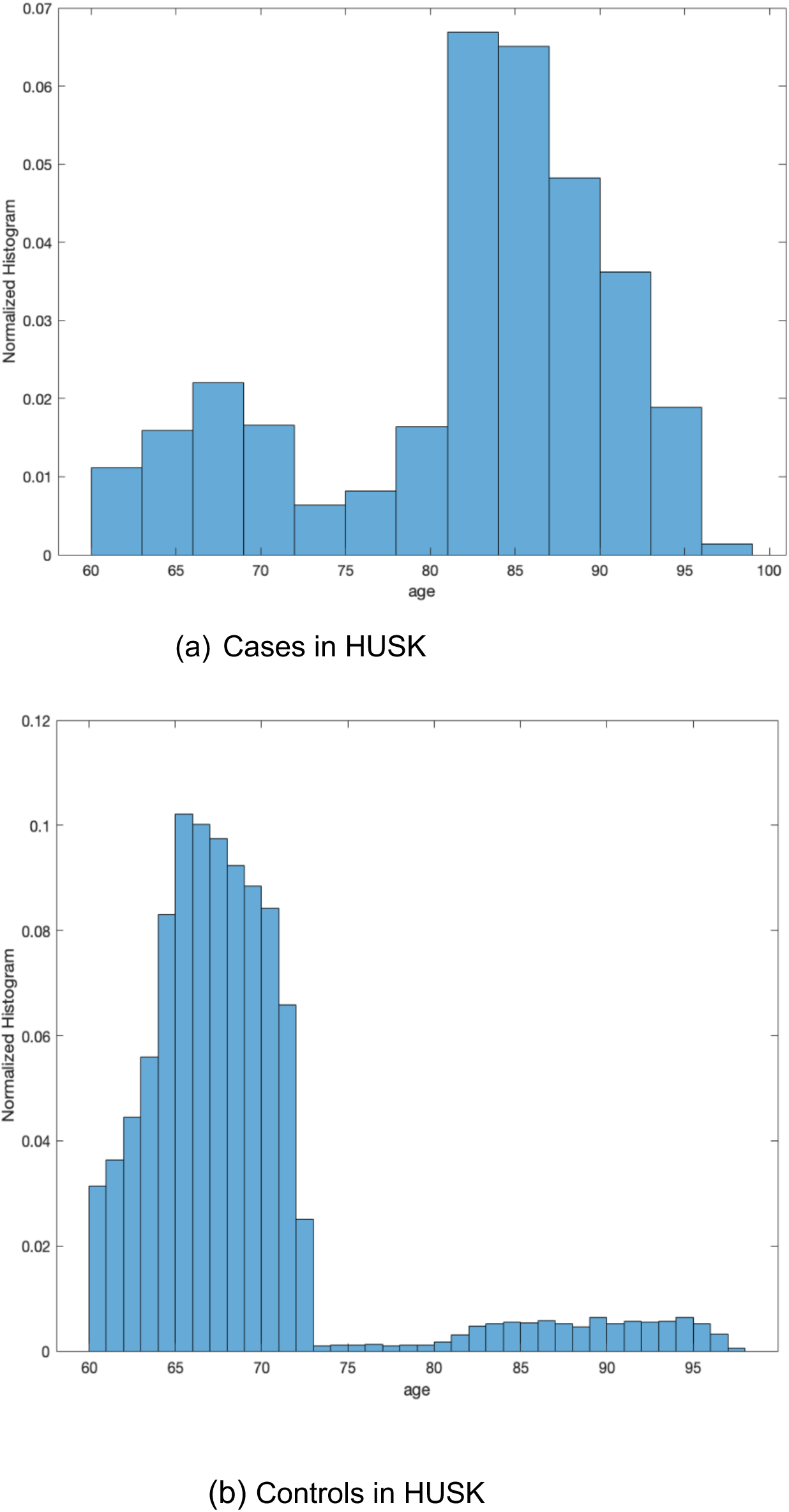
Age of onset of AD distribution for cases and age of last follow up distribution for controls for HUSK data. For cases, we used “year_of_first_diag_AD” and for controls we used the “age_last_contact” and (if available) death year (DAAR). To determine the age of onset and age of follow-up, we subtracted these dates from the year of birth and obtained the resultant ages. The dataset has biphasic distribution since nearly all participants invited in 1992-93 were born in 1925-27 and 1950-52. The participants invited in 1997-99 were born in 1953-57.

**Fig S4.**
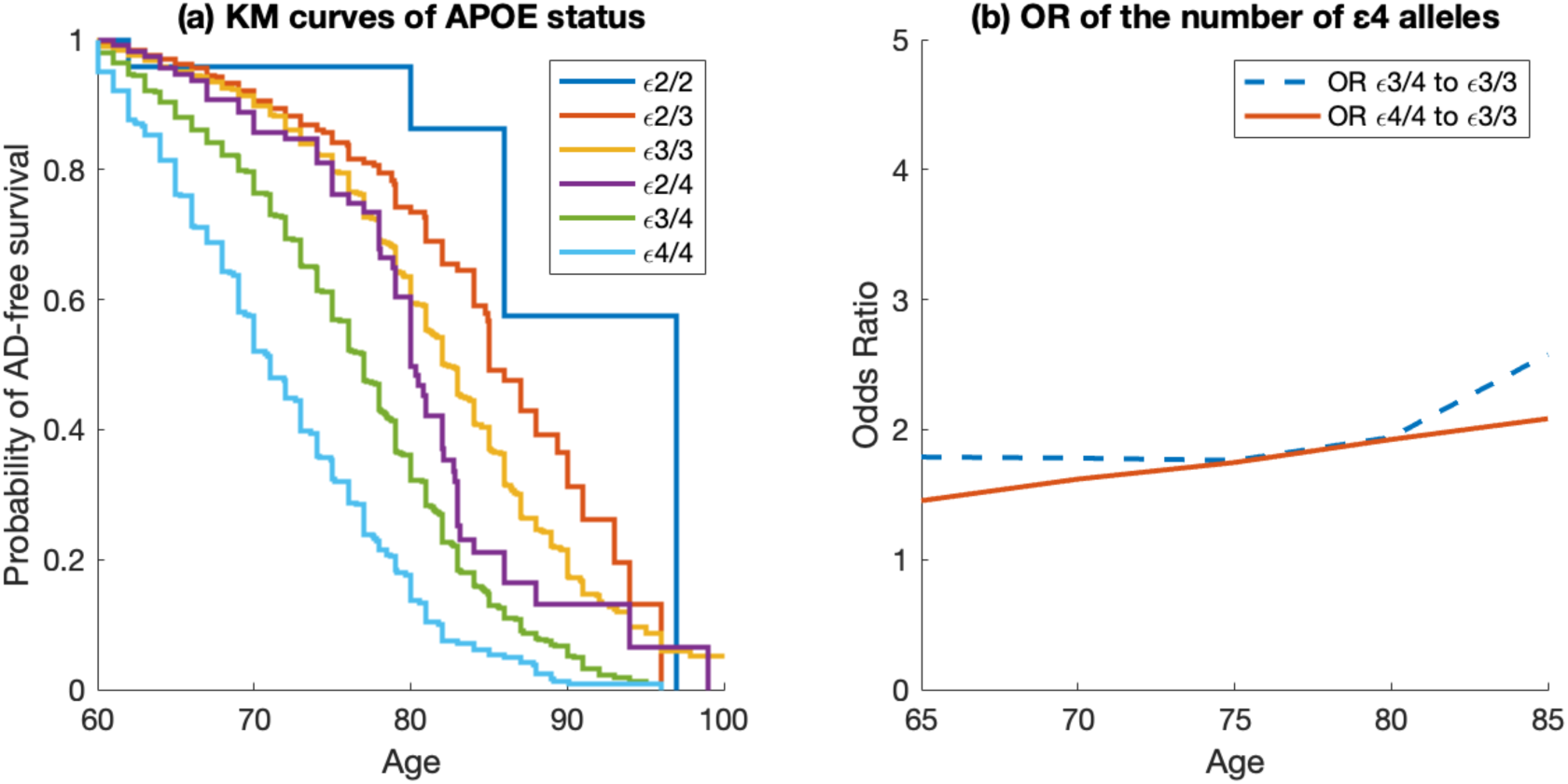
Examination of effect of *APOE* status on AD using all EADB data (a) Kaplan-Meier (KM) curves of the samples using their *APOE* status, (b) Odds Ratio (OR) of the samples with respect to the number of ε4 alleles.

**Table S1a.** OR metrics (and standard deviation) for top 5 percentile to bottom 20 percentile for EADB validation cohort.

| Predictor | Dataset | OR<sub>95/20</sub><br>at 65 | OR<sub>95/20</sub> at<br>70 | OR<sub>95/20</sub><br>at 75 | OR<sub>95/20</sub><br>at 80 | OR<sub>95/20</sub><br>at 85 |
| --- | --- | --- | --- | --- | --- | --- |
| EADB720 | all | 11.32<br>(2.24) | 14.37<br>(2.36) | 17.77<br>(2.71) | 22.10<br>(5.18) | 40.25<br>(19.5) |
| Desikan | all | 7.21<br>(1.34) | 8.95<br>(1.42) | 10.34<br>(1.98) | 12.90<br>(2.34) | 25.44<br>(13.63) |
| EADB720 | e3e3 | 1.60<br>(0.57) | 2.06<br>(0.63) | 2.43<br>(0.88) | 2.88<br>(0.94) | 3.94<br>(1.37) |
| Desikan | e3e3 | 1.23<br>(0.43) | 1.63<br>(0.59) | 1.84<br>(0.77) | 2.14<br>(0.88) | 2.62<br>(1.02) |

**Table S1b.**
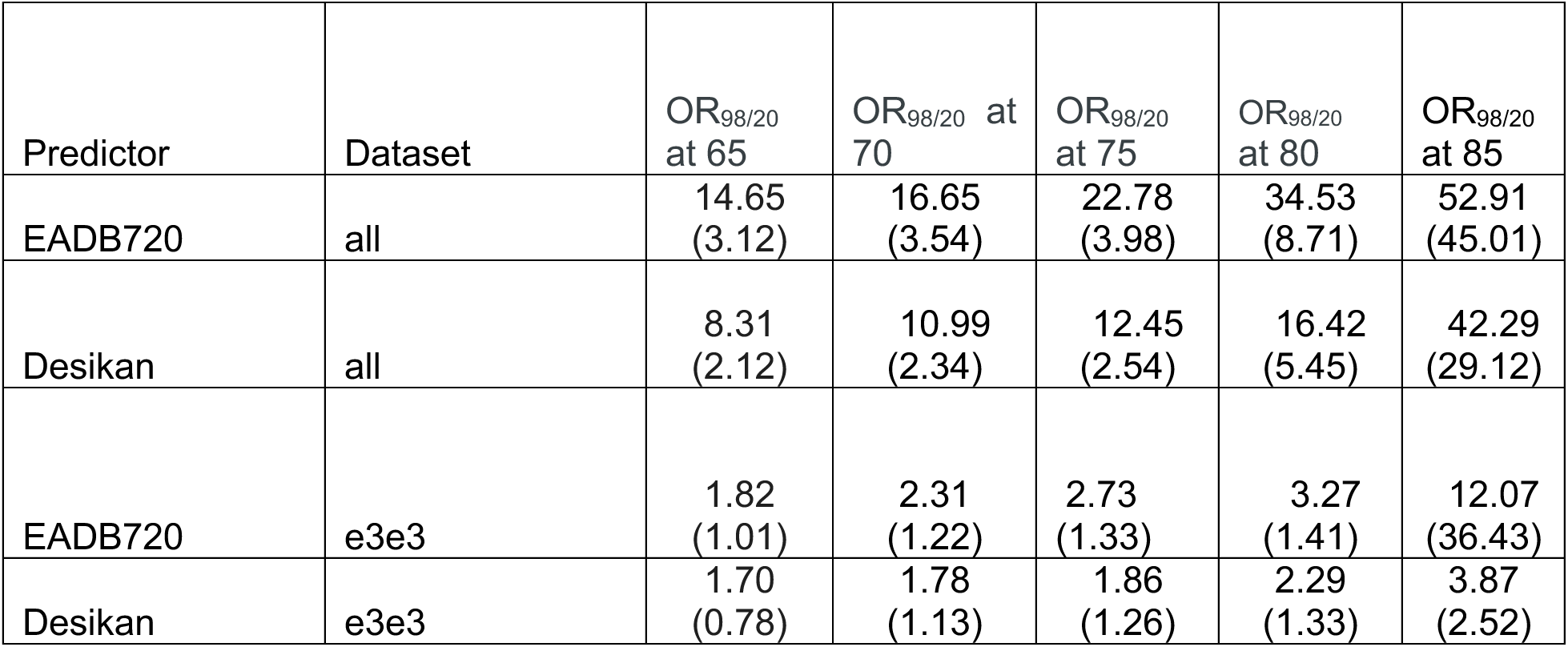
OR metrics (and standard deviation) for top 2 percentile to bottom 20 percentile for EADB validation cohort.

**Table S1c.**
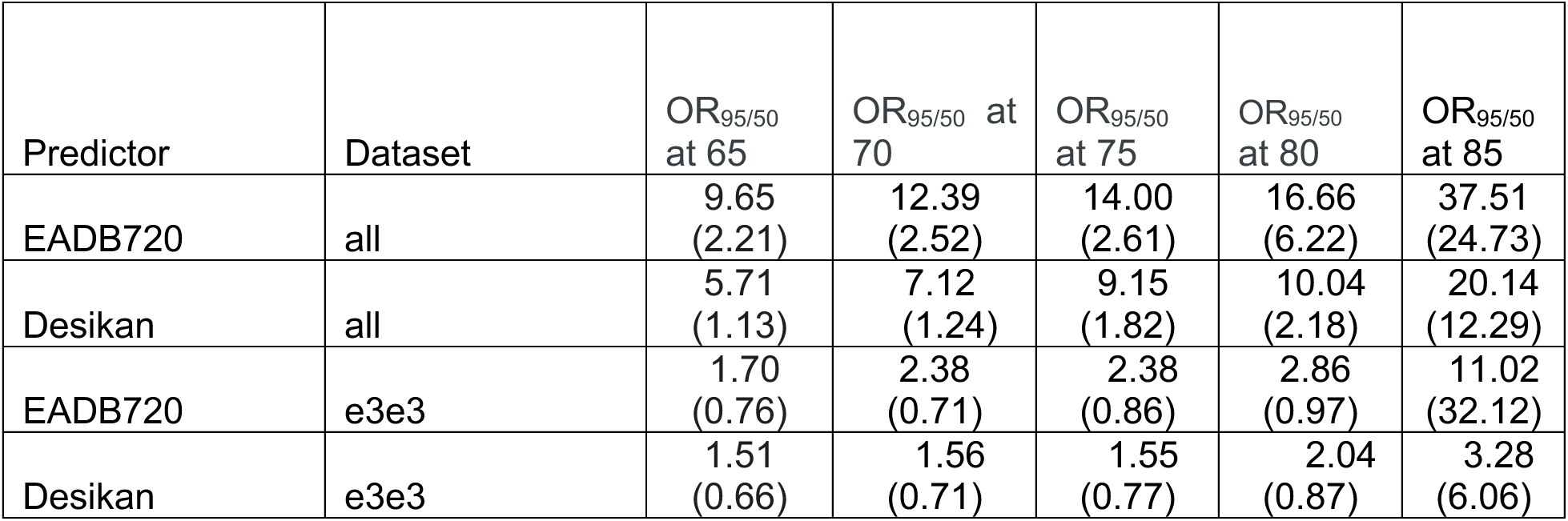
OR metrics (and standard deviation) for top 5 percentile to bottom 50 percentile for EADB validation cohort.

**Table S2a.** HUSK cohort and *APOE* status.

|  | Cases | Controls |
| --- | --- | --- |
| Total Sample (n) | 757 | 23,962 |
| Mean age of onset (cases)<br>or last follow up (controls) | 82.0 | 68.8 |
| ε2/2 percentage | 0 | 0.5 |
| ε2/3 percentage | 4.9 | 9.2 |
| ε3/3 percentage | 45.2 | 57.8 |
| ε3/4 percentage | 37.6 | 27.3 |
| ε2/4 percentage | 2.8 | 2.3 |
| ε4/4 percentage | 9.5 | 2.9 |

**Table S2b.**
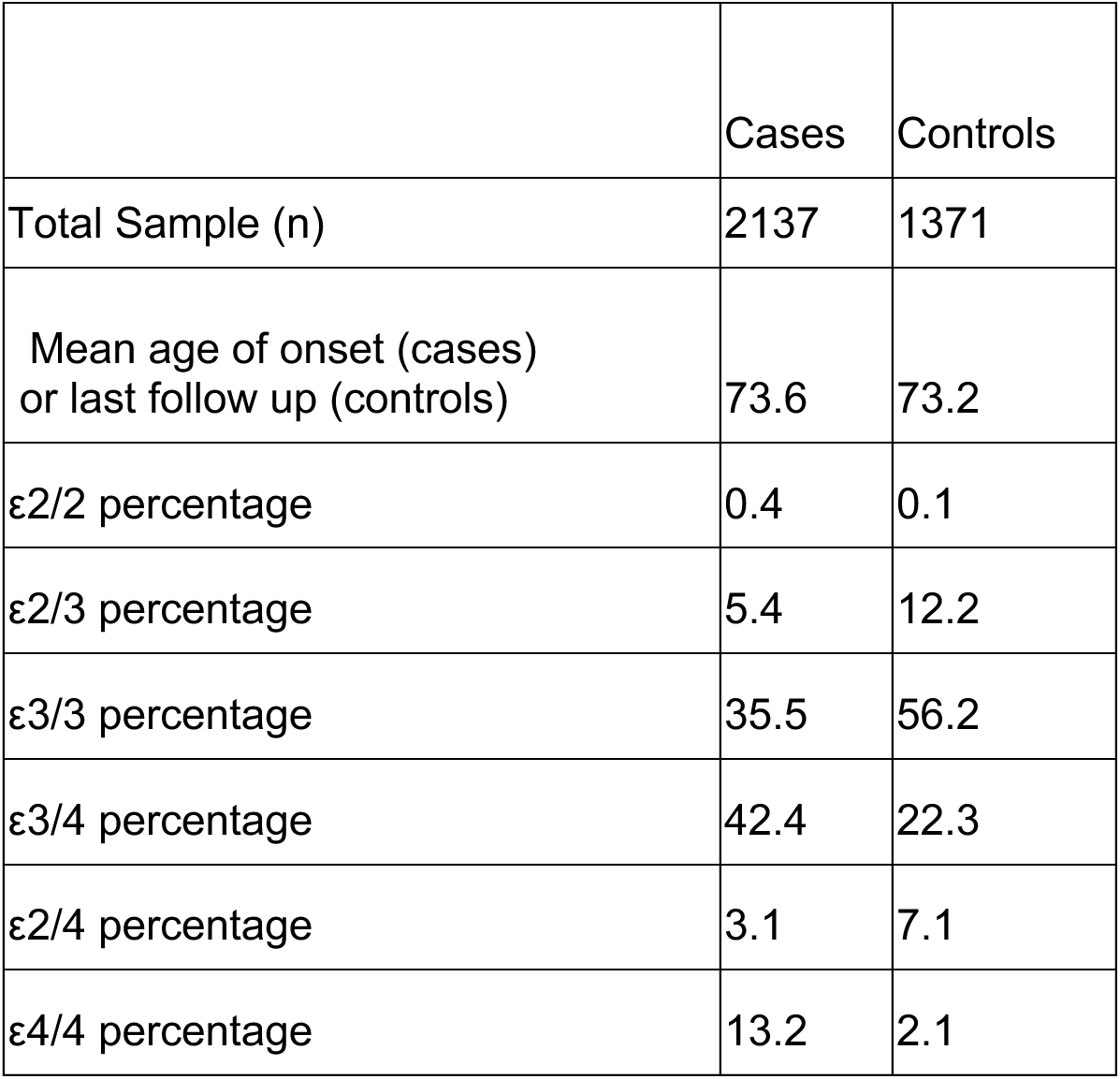
DemGene cohort and *APOE* status.

|  | Cases | Controls |
| --- | --- | --- |
| Total Sample (n) | 2137 | 1371 |
| Mean age of onset (cases)<br>or last follow up (controls) | 73.6 | 73.2 |
| ε2/2 percentage | 0.4 | 0.1 |
| ε2/3 percentage | 5.4 | 12.2 |
| ε3/3 percentage | 35.5 | 56.2 |
| ε3/4 percentage | 42.4 | 22.3 |
| ε2/4 percentage | 3.1 | 7.1 |
| ε4/4 percentage | 13.2 | 2.1 |

**Table S2c.** PHS validation in DemGene data. C-index and OR80/20 values when EADB720 is applied on all and ε3/3 individuals on DemGene data. The Desikan model’s performance is also included in the comparison.

| **DemGene**<br>**all samples** | C-index | OR<sub>80/20</sub><br>at 65 | OR<sub>80/20</sub><br>at 70 | OR<sub>80/20</sub><br>at 75 | OR<sub>80/20</sub><br>at 80 | OR<sub>80/20</sub><br>at 85 |
| --- | --- | --- | --- | --- | --- | --- |
| EADB720 | 0.67<br>(0.006) | 10.30<br>(1.01) | 5.96<br>(0.78) | 7.71<br>(0.71) | 8.02<br>(0.84) | 7.82<br>(0.92) |
| Desikan | 0.66<br>(0.006) | 8.39<br>(0.89) | 5.52<br>(0.86) | 6.60<br>(0.67) | 6.47<br>(0.99) | 7.69<br>(0.84) |
| **ε3/3**<br>**samples** |  |  |  |  |  |  |
| EADB720 | 0.55<br>(0.004) | 3.30<br>(0.91) | 1.70<br>(0.54) | 1.87<br>(0.48) | 1.62<br>(0.57) | 1.76<br>(0.53) |
| Desikan | 0.52<br>(0.011) | 1.47<br>(0.78) | 1.02<br>(0.51) | 1.07<br>(0.62) | 1.12<br>(0.87) | 1.45<br>(0.68) |

**Table S2d.** PHS validation in HUSK data using cases and controls. C-index and OR80/20 values when EADB720 is applied on all and ε3/3 individuals in the HUSK test data. The Desikan model’s performance is also included in the comparison.

| **HUSK all**<br>**samples** | C-index | OR<sub>80/20</sub><br>at 65 | OR<sub>80/20</sub><br>at 70 | OR<sub>80/20</sub><br>at 75 | OR<sub>80/20</sub><br>at 80 | OR<sub>80/20</sub><br>at 85 |
| --- | --- | --- | --- | --- | --- | --- |
| EADB720 | 0.72<br>(0.012) | 4.61<br>(0.72) | 10.44<br>(1.12) | 5.46<br>(0.81) | 7.53<br>(0.84) | 6.72<br>(0.57) |
| Desikan | 0.70<br>(0.011) | 8.39<br>(0.79) | 9.05<br>(0.65) | 6.74<br>(0.83) | 5.66<br>(0.76) | 4.58<br>(0.69) |
| **ε3/3**<br>**samples** |  |  |  |  |  |  |
| EADB720 | 0.57<br>(0.012) | 1.44<br>(0.51) | 1.86<br>(0.54) | 1.55<br>(0.72) | 1.61<br>(0.64) | 1.92<br>(0.71) |
| Desikan | 0.54<br>(0.012) | 1.47<br>(0.60) | 2.21<br>(0.66) | 1.03<br>(0.51) | 0.98<br>(0.82) | 0.89<br>(0.65) |

**Table S2e.** PHS validation in HUSK data using only cases. C-index and OR80/20 values when EADB720 is applied on all and ε3/3 individuals in the HUSK test data. The Desikan model’s performance is also included in the comparison.

| **HUSK AD cases** | C-index | OR<sub>80/20</sub><br>at 65 | OR<sub>80/20</sub><br>at 70 | OR<sub>80/20</sub><br>at 75 | OR<sub>80/20</sub><br>at 80 | OR<sub>80/20</sub><br>at 85 |
| --- | --- | --- | --- | --- | --- | --- |
| EADB720 | 0.61<br>(0.017) | 2.45<br>(0.64) | 4.64<br>(0.82) | 4.72<br>(0.76) | 5.24<br>(0.89) | 5.59<br>(0.82) |
| Desikan | 0.59<br>(0.014) | 1.75<br>(0.52) | 4.08<br>(0.69) | 4.93<br>(0.52) | 4.63<br>(0.81) | 4.03<br>(0.77) |
| **$\epsilon 3/3$ samples** |  |  |  |  |  |  |
| EADB720 | 0.54<br>(0.18) | 1.81<br>(0.59) | 1.57<br>(0.55) | 1.33<br>(0.67) | 1.45<br>(0.59) | 1.94<br>(0.79) |
| Desikan | 0.50<br>(0.019) | 1.31<br>(0.42) | 1.04<br>(0.52) | 0.66<br>(0.49) | 1.50<br>(0.59) | 1.07<br>(0.44) |

**Table S3.**
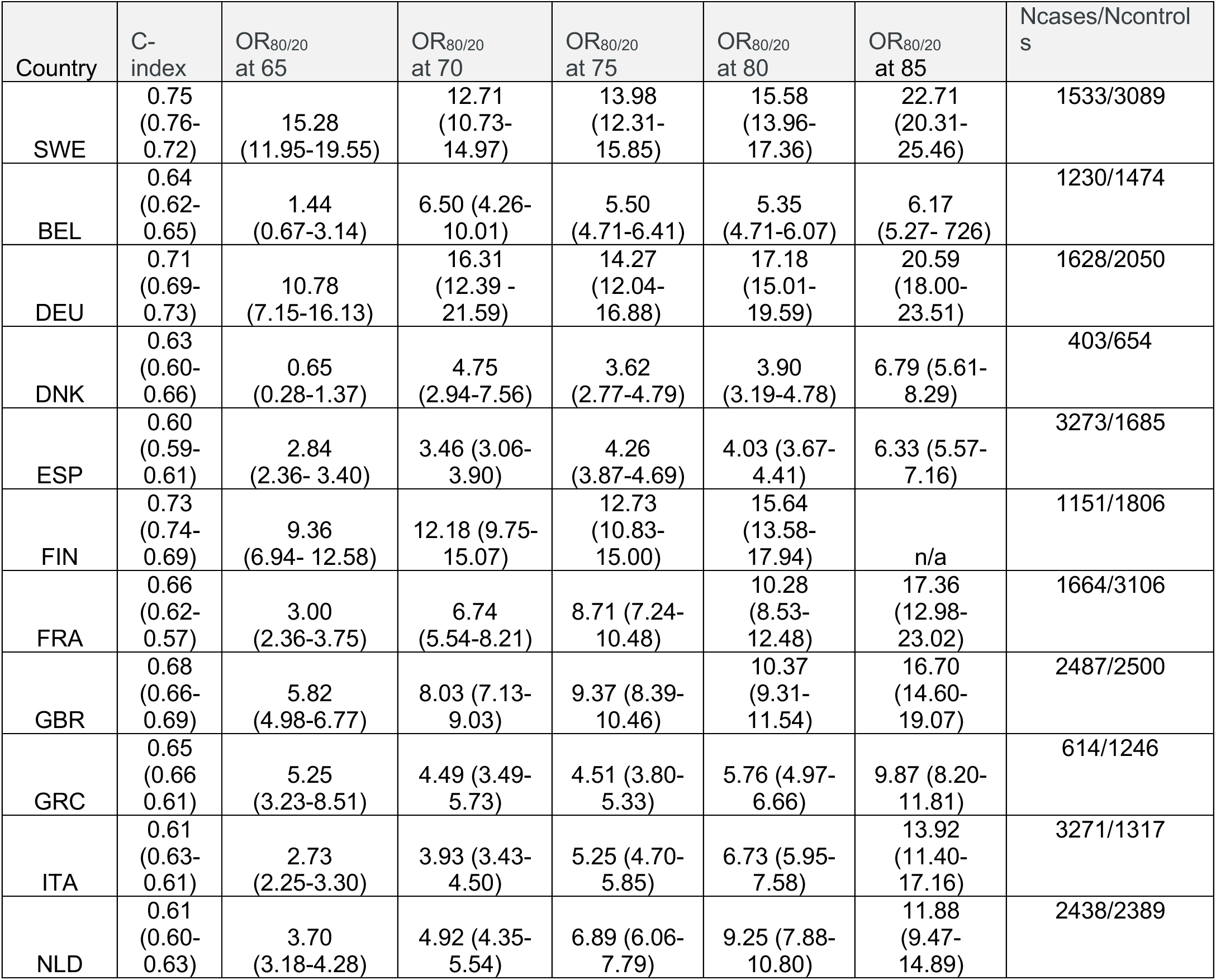
Leave one Out (LOO) analysis of the EADB PHS model for different countries. (Belgium (BEL), Denmark (DNK), Finland (FIN), France (FRA), Germany (DEU), Greece (GRC), Italy (ITA), Spain (ESP), Sweden (SWE), The Netherlands (NLD) and the Great Britain (GBR))

| Country | C-index | OR<sub>80/20</sub><br>at 65 | OR<sub>80/20</sub><br>at 70 | OR<sub>80/20</sub><br>at 75 | OR<sub>80/20</sub><br>at 80 | OR<sub>80/20</sub><br>at 85 | Ncases/Ncontrols |
| --- | --- | --- | --- | --- | --- | --- | --- |
| SWE | 0.75<br>(0.76-0.72) | 15.28<br>(11.95-19.55) | 12.71<br>(10.73-14.97) | 13.98<br>(12.31-15.85) | 15.58<br>(13.96-17.36) | 22.71<br>(20.31-25.46) | 1533/3089 |
| BEL | 0.64<br>(0.62-0.65) | 1.44<br>(0.67-3.14) | 6.50 (4.26-10.01) | 5.50<br>(4.71-6.41) | 5.35<br>(4.71-6.07) | 6.17<br>(5.27- 726) | 1230/1474 |
| DEU | 0.71<br>(0.69-0.73) | 10.78<br>(7.15-16.13) | 16.31<br>(12.39 - 21.59) | 14.27<br>(12.04-16.88) | 17.18<br>(15.01-19.59) | 20.59<br>(18.00-23.51) | 1628/2050 |
| DNK | 0.63<br>(0.60-0.66) | 0.65<br>(0.28-1.37) | 4.75<br>(2.94-7.56) | 3.62<br>(2.77-4.79) | 3.90<br>(3.19-4.78) | 6.79 (5.61-8.29) | 403/654 |
| ESP | 0.60<br>(0.59-0.61) | 2.84<br>(2.36- 3.40) | 3.46 (3.06-3.90) | 4.26<br>(3.87-4.69) | 4.03 (3.67-4.41) | 6.33 (5.57-7.16) | 3273/1685 |
| FIN | 0.73<br>(0.74-0.69) | 9.36<br>(6.94- 12.58) | 12.18 (9.75-15.07) | 12.73<br>(10.83-15.00) | 15.64<br>(13.58-17.94) | n/a | 1151/1806 |
| FRA | 0.66<br>(0.62-0.57) | 3.00<br>(2.36-3.75) | 6.74<br>(5.54-8.21) | 8.71 (7.24-10.48) | 10.28<br>(8.53-12.48) | 17.36<br>(12.98-23.02) | 1664/3106 |
| GBR | 0.68<br>(0.66-0.69) | 5.82<br>(4.98-6.77) | 8.03 (7.13-9.03) | 9.37 (8.39-10.46) | 10.37<br>(9.31-11.54) | 16.70<br>(14.60-19.07) | 2487/2500 |
| GRC | 0.65<br>(0.66-0.61) | 5.25<br>(3.23-8.51) | 4.49 (3.49-5.73) | 4.51 (3.80-5.33) | 5.76 (4.97-6.66) | 9.87 (8.20-11.81) | 614/1246 |
| ITA | 0.61<br>(0.63-0.61) | 2.73<br>(2.25-3.30) | 3.93 (3.43-4.50) | 5.25 (4.70-5.85) | 6.73 (5.95-7.58) | 13.92<br>(11.40-17.16) | 3271/1317 |
| NLD | 0.61<br>(0.60-0.63) | 3.70<br>(3.18-4.28) | 4.92 (4.35-5.54) | 6.89 (6.06-7.79) | 9.25 (7.88-10.80) | 11.88<br>(9.47-14.89) | 2438/2389 |

**Table S4.** C-index (and its standard deviation) of our sex specific PHS models when sex-specific samples were used for training and test. In other words, “sex-matched” in the first column corresponds to developing model for men by using only men and developing another model for women by using only women while the “sex-mismatched” is vice versa. Comparison is done for all samples and in subsamples with neutral *APOE* (ε3/3 samples)

| Dataset used for train<br>Full/half/sex-matched/sex-mismatched | Dataset used for validation<br>All/ $\epsilon 3\epsilon 3$ | Dataset used for validation<br>Male/female/all | c-index | std |
| --- | --- | --- | --- | --- |
| full | all | all | 0.6670 | 0.0491 |
| half | all | all | 0.6636 | 0.0486 |
| sex-matched | all | all | 0.6636 | 0.0481 |
| sex-mismatched | all | all | 0.6614 | 0.0490 |
| full | all | female | 0.6755 | 0.0571 |
| half | all | female | 0.6719 | 0.0566 |
| sex-matched | all | female | 0.6744 | 0.0559 |
| sex-mismatched | all | female | 0.6671 | 0.0585 |
| full | all | male | 0.6537 | 0.0391 |
| half | all | male | 0.6506 | 0.0401 |
| sex-matched | all | male | 0.6470 | 0.0407 |
| sex-mismatched | all | male | 0.6518 | 0.0368 |
| full | $\epsilon 3\epsilon 3$ | all | 0.5660 | 0.0183 |
| half | $\epsilon 3\epsilon 3$ | all | 0.5561 | 0.0180 |
| sex-matched | $\epsilon 3\epsilon 3$ | all | 0.5576 | 0.0222 |
| sex-mismatched | $\epsilon 3\epsilon 3$ | all | 0.5475 | 0.0178 |
| full | $\epsilon 3\epsilon 3$ | female | 0.5716 | 0.0290 |
| half | $\epsilon 3\epsilon 3$ | female | 0.5598 | 0.0204 |
| sex-matched | $\epsilon 3\epsilon 3$ | female | 0.5699 | 0.0251 |
| sex-mismatched | $\epsilon 3\epsilon 3$ | female | 0.5439 | 0.0313 |
| full | $\epsilon 3\epsilon 3$ | male | 0.5584 | 0.0223 |
| half | $\epsilon 3\epsilon 3$ | male | 0.5508 | 0.0301 |
| sex-matched | $\epsilon 3\epsilon 3$ | male | 0.5407 | 0.0360 |
| sex-mismatched | $\epsilon 3\epsilon 3$ | male | 0.5515 | 0.0212 |

**Table S5.** Comparison of the performance of variants used in EADB model when our determined effect sizes were used using our PHS development procedure vs when the effect sizes of the EADB GWAS in [21] were used. The comparison is done for ε3/3 samples.

| $\epsilon 3/3$<br>samples | C-index | OR<sub>80/20</sub><br>at 65 | OR<sub>80/20</sub><br>at 70 | OR<sub>80/20</sub><br>at 75 | OR<sub>80/20</sub><br>at 80 | OR<sub>80/20</sub><br>at 85 |
| --- | --- | --- | --- | --- | --- | --- |
| EADB720 | 0.57<br>(0.009) | 1.95<br>(0.49) | 2.32<br>(0.40) | 2.77<br>(0.44) | 3.51<br>(0.48) | 3.52<br>(0.87) |
| SNPs in<br>EADB720<br>with EADB<br>GWAS<br>effect size | 0.54<br>(0.01) | 1.27<br>(0.36) | 1.65<br>(0.28) | 1.47<br>(0.26) | 1.68<br>(0.27) | 2.23<br>(0.42) |
| Desikan | 0.54<br>(0.008) | 1.27<br>(0.04) | 1.50<br>(0.56) | 1.82<br>(0.48) | 1.87<br>(0.26) | 2.00<br>(0.27) |

**Table S6.** Comparison of the performance of EADB720 and other models for HUSK and DemGene datasets. The comparison is done for ε3/3 samples. EADB720 outperforms PGC GWAS (pval=0.01 for HUSK, pval=0.002 for DemGene), Meta Analysis (pval=0.02 for HUSK, pval=0.005 for DemGene) and EADB PRS (pval=0.003 for HUSK, pval=0.002 for DemGene)

| HUSK<br>$\epsilon 3/3$<br>samples | C-index |
| --- | --- |
| EADB720 | 0.57<br>(0.012) |
| Desikan | 0.54<br>(0.012) |
| PGC GWAS | 0.55<br>(0.014) |
| Meta Analysis | 0.55<br>(0.016) |
| EADB PRS | 0.54<br>(0.010) |

| DemGene<br>$\epsilon 3/3$<br>samples | C-index |
| --- | --- |
| EADB720 | 0.55<br>(0.004) |
| Desikan | 0.52<br>(0.011) |
| PGC GWAS | 0.52<br>(0.012) |
| Meta Analysis | 0.53<br>(0.015) |
| EADB PRS | 0.53<br>(0.007) |

## Supplementary Authors (in arbitrary order according to cohorts)

Sebastiaan Engelborghs 1, 2, 3, Rik Vandenberghe 4, 5, Camille Charbonnier 6, David Wallon 7, Giuliano Binetti 8, Silvia Fostinelli 8, Sonia Bellini 9, Chiara Fenoglio 10, 11, Andrea Arighi 12, Maria Serpente 12, Valerio Napolioni 13, Federico Paolini Paoletti 14, Lorenzo Gaetani 15, Caterina Chillotti 16, Claudia Pisanu 17, Elisa Conti 18, Ildebrando Appollonio 18, 19, Carlo Ferrarese 18, 19, Elisa Rubino 20, Fausto Roveta 20, Michela Orsini 21, Maria Gabriella Vita 22, Guido Maria Giuffrè 22, 23, Nerisa Banaj 24, Filomena Iannuzzi 25, Alessandra Lauria 26, Fabrizio Tagliavini 27, Johannes Kornhuber 28, 126, Gabor C. Petzold 29, 30, Michael T. Heneka 29, 31, Christoph Laske 32, 33, Robert Perneczky 34, 35, 36, 37, Josef Priller 38, 39, Annika Spottke 29, 40, Stefan Teipel 41, 42, Michael Ewers 43, Rainer Malik 43, Lutz Frölich 44, 119, Annette M. Hartmann 45, 46, Oliver Goldhardt 47, Matthias Schmid 29, 48, Per Hoffmann 49, 50, Markus M Nöthen 49, 130, Kayenat Parveen 51, 52, Victor Andrade 51, 52, Peter Riederer 53, Peter Fischer 54, Christopher Clark 55, Martin J. Herrmann 56, Thomas Polak 56, Heike Weber 56, Michael Wagner 29, 31, Mary Yannakoulia 57, Georgios Hadjigeorgiou 58, Costas Anastasiou 57, Kalina Yonkova Mihova 59, Ignacio Alvarez 60, 83, 84, Ingrid T. Medbøen 61, 62, Karin Persson 61, 62, Gøril Rolfseng Grøntvedt 63, Lasse Pihlstrøm 64, Ole Kristian Drange 65, 66, Sigrid Botne Sando 63, 67, Kaja Nordengen 68, Marta Marquié 70, 88, 89, Claudia Olive 71, 88, Pablo García-González 71, 88, Amanda Cano 88, Raquel Puerta 72, 88, Laura Montrreal 88, Sergi Valero 70, 88, 89, Oscar Sotolongo-Grau 88, Ana Espinosa 70, 88, 89, Angela Sanabria 70, 88, 89, Gemma Ortega 70, 88, 89, Maitee Rosende-Roca 70, 88, 89, Montserrat Alegret 70, 88, 89, Lluís Tárraga 70, 88, 89, Sebastián García-Madrona 73, Anaïs Corma-Gómez 74, Juan Antonio Pineda 74, Juan Macías 74, Laura Muñoz-Delgado 75, 76, María Bernal Sánchez-Arjona 77, María Teresa Periñán 75, 76, 78, Silvia Jesús 75, 76, Alfonso Arias Pastor 79, 80, Arturo Corbatón-Anchuelo 81, 82, María Teresa Martínez-Larrad 81,82, Paz De la Guía 85, Silvia Mendoza 86, Ernesto García-Roldán 77, Miguel Martín-Bórnez 75, 76, Mercè Boada 87, 88, Asunción Lafuente 88, Inés Quintela 90, Marta Ibarria 88, Nuria Aguilera 88, Olalla Maroñas 90, Pilar Cañabate 88, Silvia Preckler 88, Susana Diego 88, Celia Painous 91, Manuel Fernandez 91, 92, 93, Yaroslau Compta 91, Consuelo Cháfer-Pericás 94, Lourdes Álvarez-Sánchez 94, Fernando Cardona 92, 95, Juan Marín-Muñoz 96, Ana Belén Pastor 97, 98, Teodoro del Ser 99, Manuel Menéndez-González 100, 101, 102, Alberto Lleó 92, 103, Daniel Alcolea 92, 103, Jaime Kulisevsky 92, 104, Javier Pagonabarraga 92, 104, Olivia Belbin 92, 103, Carmen Lage 92, 105, Sara López-García 92, 106, Catherine Bresner 107, Gill Windle 108, 109, Catherine MacLeod 110, Bob Woods 108, Simon Mead 111, 129, Keeley Brookes 112, Jonathan M Schott 113, Nick C. Fox 113, 118, Nicholas J Bass 114, Carlos Cruchaga 115, 116, Martin Dichgans 36, 117, Alison M Goate 120, John Hardy 121, Reinhard Heun 122, Michael Hüll 123, Karl-Heinz Jöckel 124, Janet A Johnston 125, Gill Livingston 114, Michelle K Lupton 127, 128, Bernadette McGuinness 125, Andrew McQuillin 114, Peter Passmore 125, Petra Proitsi 78, 127, Martin Rossor 118, Dulcenombre Gómez Garre 81, 131

1. Neuroprotection and Neuromodulation (NEUR) Research Group, Center for Neurosciences (C4N), Vrije, Universiteit Brussel (VUB), Brussel, Belgium
2. Department of Neurology, Universitair Ziekenhuis Brussel (UZ Brussel), Brussel, Belgium
3. Department of Biomedical Sciences, University of Antwerp, Antwerp, Belgium
4. Department of Neurology, UZ Leuven, Louvain, Belgium
5. Laboratory for Cognitive Neurology, Department of Neurosciences, KU Leuven, Leuven Brain Institute, Louvain, Belgium.
6. Univ Rouen Normandie, Normandie Univ, Inserm U1245 and CHU Rouen, Department of Biostatistics and CNRMAJ, F-76000 Rouen, France
7. Univ Rouen Normandie, Normandie Univ, Inserm U1245 and CHU Rouen, Department of Neurology and CNRMAJ, F-76000 Rouen, France
8. MAC-Memory Clinic and Molecular Markers Laboratory, IRCCS Istituto Centro San Giovanni di Dio Fatebenefratelli, Brescia, Italy
9. Molecular Markers Laboratory, IRCCS Istituto Centro San Giovanni di Dio Fatebenefratelli, Brescia, Italy
10. Dept. of Biomedical, Surgical and Dental Sciences, University of Milan, Milan, Italy
11. Neurodegenerative Diseases Unit, Fondazione IRCCS Ca’ Granda, Ospedale Policlinico, Milan, Italy
12. Neurodegenerative Diseases Unit, Fondazione IRCCS Ca’ Granda, Ospedale Maggiore Policlinico, Milan, Italy
13. School of Biosciences and Veterinary Medicine, University of Camerino, Camerino, Italy
14. Centre for Memory Disturbances, Lab of Clinical Neurochemistry, Section of Neurology, University of Perugia, Perugia, Italy
15. Lab of Clinical Neurochemistry, Section of Neurology, University of Perugia, Perugia, Italy
16. Unit of Clinical Pharmacology, University Hospital Agency of Cagliari, Cagliari, Italy
17. Department of Biomedical Sciences, Section of Neuroscience and Clinical Pharmacology,University of Cagliari, Cagliari, Italy
18. School of Medicine and Surgery, University of Milano-Bicocca, Monza, Italy
19. Neurology Unit, IRCCS “San Gerardo dei Tintori”, Monza, Italy
20. Department of Neuroscience “Rita Levi Montalcini”, University of Torino, Torino, Italy
21. Psychology Unit, IRCCS Fondazione Policlinico Universitario A. Gemelli, Rome, Italy
22. Neurology Unit, IRCCS Fondazione Policlinico Universitario A. Gemelli, Rome, Italy
23. Department of Neuroscience, Università Cattolica del Sacro Cuore, Rome, Italy
24. Laboratory of Neuropsychiatry, Clinical Neuroscience and Neurorehabilitation Department, IRCCS Santa Lucia Foundation, Rome, Italy
25. Laboratory of Experimental Neuropsychobiology, Clinical Neuroscience and Neurorehabilitation Department, IRCCS Santa Lucia Foundation, Rome, Italy
26. High Intensity Neurorehabilitation Unit, Fondazione Policlinico Universitario ’A. Gemelli’ IRCCS, Rome, Italy
27. Fondazione IRCCS Istituto Neurologico Carlo Besta, Milan, Italy
28. Department of Psychiatry and Psychotherapy, Universitätsklinikum Erlangen, and Friedrich-Alexander Universität Erlangen-Nürnberg, Erlangen, Germany.
29. German Center for Neurodegenerative Diseases (DZNE), Bonn, Germany
30. Department of Vascular Neurology, University Hospital Bonn, Bonn, Germany
31. Department for Neurodegenerative Diseases and Geriatric Psychiatry, University Hospital Bonn, Venusberg-Campus 1, 53127 Bonn, Germany
32. German Center for Neurodegenerative Diseases (DZNE), Tübingen, Germany
33. Section for Dementia Research, Hertie Institute for Clinical Brain Research and Department of Psychiatry and Psychotherapy, University of Tübingen, Tübingen, Germany
34. Ageing Epidemiology Research Unit (AGE), School of Public Health, Imperial College London, London, UK
35. Department of Psychiatry and Psychotherapy, University Hospital, LMU Munich, Munich, Germany
36. German Center for Neurodegenerative Diseases (DZNE, Munich), Feodor-Lynen-Strasse 17, 81377 Munich, Germany
37. Munich Cluster for Systems Neurology (SyNergy) Munich, Munich, Germany
38. German Center for Neurodegenerative Diseases (DZNE), Berlin, Germany
39. Department of Psychiatry and Psychotherapy, Charité, Charitéplatz 1, 10117 Berlin, Germany
40. Department of Neurology, University of Bonn, Venusberg-Campus 1, 53127 Bonn, Germany
41. Department of Psychosomatic Medicine, Rostock University Medical Center, Gehlsheimer Str. 20, 18147 Rostock
42. German Center for Neurodegenerative Diseases (DZNE), Rostock, Germany
43. Institute for Stroke and Dementia Research (ISD),University Hospital, Ludwig-Maximilian University Munich
44. Department of Geriatric Psychiatry, Central Institute for Mental Health Mannheim, Faculty Mannheim, University of Heidelberg, Germany
45. Comprehensive Center for Clinical Neurosciences and Mental Health, Medical University of Vienna, Vienna, Austria
46. Department of Psychiatry and Psychotherapy, Medical University of Vienna, Vienna, Austria
47. Center for Cognitive Disorders, Department of Psychiatry and Psychotherapy, TUM University Hospital, Technical University of Munich, School of Medicine, Munich, Germany
48. Institute of Medical Biometry, Informatics and Epidemiology, University Hospital of Bonn, Bonn, Germany.
49. Institute of Human Genetics; University of Bonn, School of Medicine & University Hospital Bonn, 53127 Bonn, Germany
50. Institute of Medical Genetics and Pathology, University Hospital Basel, Basel, Switzerland.
51. Department of Neurodegeneration and Geriatric Psychiatry, University of Bonn, 53127 Bonn, Germany.
52. Division of Neurogenetics and Molecular Psychiatry, Department of Psychiatry and Psychotherapy, University of Cologne, Medical Faculty, 50937 Cologne, Germany.
53. Center of Mental Health, Clinic and Policlinic of Psychiatry, Psychosomatics and Psychotherapy, University Hospital of Würzburg, Würzburg, Germany
54. Department of Psychiatry, Social Medicine Center East-Donauspital, Vienna, Austria
55. Institute for Regenerative Medicine, University of Zürich, Switzerland
56. Department of Psychiatry, Psychosomatics and Psychotherapy, Center of Mental Health, University Hospital of Würzburg
57. Department of Nutrition and Diatetics, Harokopio University, Athens, Greece
58. Department of Neurology, Medical School, University of Cyprus, Cyprus
59. Molecular Medicine Center, Department of Medical chemistry and biochemistry, Medical University of Sofia, Bulgaria
60. Neurosciences, Germans Trias I Pujol Research Institute (IGTP), Badalona, Barcelona, Spain
61. Department of Geriatric Medicine, Oslo University Hospital, Oslo, Norway
62. Norwegian National Centre for Ageing and Health, Vestfold Hospital Trust, Tønsberg, Norway
63. Department of Neuromedicine and Movement Science, Faculty of Medicine and Health Sciences, Norwegian University of Science and Technology (NTNU), Trondheim, Norway
64. Department of Neurology, Oslo University Hospital, Oslo, Norway
65. Centre for Precision Psychiatry, Division of Mental Health and Addiction, Oslo University Hospital
66. Department of Psychiatry, Sørlandet hospital HF, Arendal/Kristiansand, Norway
67. Department of Neurology and Clinical Neurophysiology, St. Olavs Hospital, Trondheim University Hospital, Trondheim, Norway
68. Department of Neurology, Akershus University Hospital, Lørenskog Norway
69. Ace Alzheimer Center Barcelona, Universitat Internacional de Catalunya (UIC), Barcelona, Spain
70. Networking Research Center on Neurodegenerative Diseases (CIBERNED), Instituto de Salud Carlos III, Madrid, Spain
71. PhD Program in Biotechnology, Faculty of Pharmacy and Food Sciences, University of Barcelona, 08028 Barcelona, Spain
72. PhD Program in Biotechnology, Faculty of Pharmacy and Food Sciences, University of Barcelona, 08028 Barcelona, Spain
73. Hospital Universitario Ramon y Cajal, IRYCIS, Madrid
74. Departamento de Medicina, Hospital Universitario Virgen de Valme. Universidad de Sevilla
75. Centro de Investigación Biomédica en Red sobre Enfermedades Neurodegenerativas, Instituto de Salud Carlos III, Madrid, Spain
76. Unidad de Trastornos del Movimiento, Servicio de Neurología, Instituto de Biomedicina de Sevilla, Hospital Universitario Virgen del Rocío/CSIC/Universidad de Sevilla, Seville, Spain
77. Unidad de Demencias, Servicio de Neurología. Instituto de Biomedicina de Sevilla (IBiS), Hospital Universitario Virgen del Rocío/CSIC/Universidad de Sevilla, Seville, Spain
78. Centre for Preventive Neurology, Wolfson Institute of Population Health, Queen Mary University of London, London, United Kingdom.
79. Institut de Recerca Biomedica de Lleida (IRBLLeida), Lleida, Spain
80. Unitat Trastorns Cognitius, Hospital Universitari Santa Maria de Lleida, Lleida, Spain
81. Instituto de Investigación Sanitaria, Hospital Clínico San Carlos (IdISSC), Madrid, Spain
82. Spanish Biomedical Research Centre in Diabetes and Associated Metabolic Disorders(CIBERDEM), Madrid, Spain
83. Fundació Docència i Recerca MútuaTerrassa, Terrassa, Barcelona, Spain
84. Memory Disorders Unit, Department of Neurology, Hospital Universitari Mutua de Terrassa, Terrassa, Barcelona, Spain
85. Instituto Andaluz de Neurociencia
86. Alzheimer Research Center & Memory Clinic, Instituto Andaluz de Neurociencia, Málaga, Spain.
87. CIBERNED, Network Center for Biomedical Research in Neurodegenerative Diseases, National Institute of Health Carlos III, Madrid, Spain
88. Ace Alzheimer Center Barcelona, Universitat Internacional de Catalunya (UIC), Barcelona, Spain.
89. Networking Research Center on Neurodegenerative Diseases (CIBERNED), Instituto de Salud Carlos III, Madrid, Spain.
90. Grupo de Medicina Xenómica, Fundación Pública Galega de Medicina Xenómica, Santiago de Compostela, Spain.
91. Parkinson’s Disease & Movement Disorders Unit, Neurology Service, Hospital Clínic I Universitari de Barcelona; IDIBAPS, CIBERNED (CB06/05/0018-ISCIII), ERN-RND, Institut Clínic de Neurociències (Maria de Maeztu Excellence Centre), Universitat de Barcelona. Barcelona, Catalonia, Spain
92. CIBERNED, Network Center for Biomedical Research in Neurodegenerative Diseases, National Institute of Health Carlos III, Madrid, Spain
93. Universitat de Barcelona (UB)
94. Instituto de Investigación Sanitaria La Fe, Hospital Universitari i Politècnic La Fe, Valencia, Spain
95. Unitat de Genètica Molecular, Institut de Biomedicina de València-CSIC, Valencia, Spain
96. Unidad de Demencias. Hospital Clínico Universitario Virgen de la Arrixaca, Palma, Spain
97. BT-CIEN
98. CIEN Foundation/Queen Sofia Foundation Alzheimer Center
99. Department of Neurology/CIEN Foundation/Queen Sofia Foundation Alzheimer Center
100. Departamento de Medicina, Universidad de Oviedo, Oviedo, Spain
101. Instituto de Investigación Sanitaria del Principado de Asturias (ISPA)
102. Servicio de Neurología. Hospital Universitario Central de Asturias, Oviedo, Spain
103. Sant Pau Memory Unit, IR SANT PAU, Hospital de la Santa Creu i Sant Pau, Barcelona, Spain
104. Movement Disorders Unit, Department of Neurology, Institut de Recerca de Sant Pau; Hospital de Sant Pau, Barcelona, Spain
105. Neurology Service, Marqués de Valdecilla University Hospital (University of Cantabria and IDIVAL), Santander, Spain.
106. Service of Neurology, University Hospital Marqués de Valdecilla, IDIVAL, University of Cantabria, Santander, Spain
107. Division of Psychological Medicine and Clinical Neuroscience, School of Medicine, Cardiff University, Cardiff, UK
108. School of Health Sciences, Bangor University, UK
109. Wales Centre for Ageing & Dementia Research
110. Usher Institute, The University of Edinburgh, UK
111. MRC Prion Unit at UCL, UCL Institute of Prion Diseases, London, W1W 7FF
112. Interdisciplinary Biomedical Research Center; Biosciences, Nottingham Trent University, Nottingham UK
113. Dementia Research Centre, UCL, London, WC1N 3BG
114. Division of Psychiatry, University College London, UK
115. Department of Psychiatry, Washington University School of Medicine, St. Louis Missouri, USA
116. Hope Center Program on Protein Aggregation and Neurodegeneration, Washington University School of Medicine, St. Louis, Missouri, USA
117. Institute for Stroke and Dementia Research, Klinikum der Universität München, Munich, Germany
118. Dementia Research Centre, Department of Neurodegenerative Disease, UCL Institute of Neurology, London, UK
119. Central Institute of Mental Health, Medical Faculty Mannheim, University of Heidelberg, Germany.
120. Icahn School of Medicine at Mount Sinai, New York, NY, USA
121. Department of Molecular Neuroscience, UCL, Institute of Neurology, London, UK
122. Department of Psychiatry and Psychotherapy, University of Bonn, 53127, Bonn, Germany
123. Department of Psychiatry, University of Freiburg, Freiburg, Germany (M.H.)
124. Institute for Medical Informatics, Biometry and Epidemiology, University Hospital of Essen, University Duisburg-Essen, Hufelandstr. 55, D-45147 Essen, Germany.
125. Centre for Public Health, School of Medicine, Dentistry and Biomedical Sciences, Queens University, Belfast, UK
126. Department of Psychiatry and Psychotherapy, University of Erlangen-Nuremberg, Germany
127. Department of Basic and Clinical Neuroscience, Institute of Psychiatry, Psychology and Neuroscience, Kings College London, London UK; 44
128. Brain and Mental Health, QIMR Berghofer, Herston, Queensland, Australia
129. MRC Prion Unit at UCL, Institute of Prion Diseases, London
130. Institute of Human Genetics, Department of Genomics, Life and Brain Center, University of Bonn, Bonn, Germany
131. Biomedical Research Centre in Cardiovascular Diseases(CIBERCV), Madrid, Spain

